# Elevated BrainAGE precedes cognitive impairment and improves prediction of future cognitive decline

**DOI:** 10.64898/2026.07.15.26358150

**Authors:** Elaheh Moradi, Robert Dahnke, Christian Gaser, Toni Rikkonen, Heikki Kröger, Sami Väänänen, Alina Solomon, Reijo Sund, Jussi Tohka, the Alzheimer’s Disease Neuroimaging Initiative

**Affiliations:** A.I. Virtanen Institute for Molecular Sciences, University of Eastern Finland, Kuopio 70150, Finland; Structural Brain Mapping Group, Department of Neurology, Jena University Hospital, Jena, Germany; Department of Psychiatry and Psychotherapy, Jena University Hospital, Jena, Germany; Institute of Clinical Medicine, University of Eastern Finland, Kuopio, Finland; Department of Orthopaedics, Traumatology and Hand Surgery, Kuopio University Hospital, Kuopio, Finland; Department of Clinical Radiology, Kuopio University Hospital, Kuopio, Finland; Department of Technical Physics, University of Eastern Finland, Kuopio, Finland; Division of Clinical Geriatrics, Center for Alzheimer Research, Karolinska Institute, Sweden; Ageing Epidemiology Research Unit, School of Public Health, Imperial College London, United Kingdom; Clinical Research Centre, Kuopio University Hospital, Kuopio, Finland

**Author notes:** Data used in preparation of this article were obtained from the Alzheimer’s Disease Neuroimaging Initiative (ADNI) database (adni.loni.usc.edu). As such, the investigators within the ADNI contributed to the design and implementation of ADNI and/or provided data but did not participate in analysis or writing of this report. A complete listing of ADNI investigators can be found at: http://adni.loni.usc.edu/wp-content/uploads/how_to_apply/ADNI_Acknowledgement_List.pdf.

**Keywords:** Longitudinal trajectories, Machine learning, Progression to mild cognitive impairment, Memory decline, Magnetic resonance imaging, Risk stratification

## Abstract

Magnetic Resonance Imaging (MRI) derived brain age varies substantially between individuals, but it remains unclear whether early deviations from normal brain ageing precede future cognitive decline and whether they provide predictive value beyond conventional MRI measures.

Here, we investigated whether MRI-derived brain age gap estimation (BrainAGE) identifies early structural brain ageing differences among cognitively normal individuals who later develop mild cognitive impairment (MCI) or dementia. We analysed longitudinal structural MRI data from the Alzheimer’s Disease Neuroimaging Initiative (ADNI) and replicated the main findings in the population-based Kuopio Osteoporosis Risk Factor and Prevention Study (OSTPRE).

Individuals who later converted to MCI or dementia had higher BrainAGE values several years before diagnosis and, in ADNI, showed steeper longitudinal increases than stable individuals. Elevated BrainAGE values were also associated with increased risk of future conversion to MCI in cognitively healthy individuals and faster subsequent memory decline. Cross-sectional differences and the association between BrainAGE and risk of future conversion were replicated in OSTPRE. Importantly, adding BrainAGE to models including demographic, APOE4, cognitive, and MRI-derived measures consistently improved prediction of future cognitive outcomes, with the greatest benefit observed for individuals who converted after longer follow-up. These findings show that structural brain ageing begins to diverge years before the onset of MCI. BrainAGE captures this early divergence, providing complementary information beyond conventional structural MRI measures that may improve the early identification of cognitively normal individuals at increased risk of future cognitive decline when integrated with other biomarkers.

## Introduction

Ageing is a heterogeneous biological process, and individuals differ markedly in how their brains change over time.^1, 2^ Although chronological age is a major determinant of cognitive and functional decline, people of the same age often show marked variability in brain structure, cognitive performance, and vulnerability to impairment.^3, 4, 5, 6, 7^ Such variability likely reflects the combined influence of normal ageing processes and early pathological changes, which may emerge gradually and follow partially overlapping trajectories. Quantifying how individuals diverge in their patterns of brain ageing may help explain differences in cognitive trajectories, including why some individuals remain cognitively healthy while others progress toward decline.

Neuroimaging-based estimates of brain age provide a way to quantify this heterogeneity at the individual level. Brain age gap estimation (BrainAGE) captures the deviation between a person’s predicted brain age and their chronological age.^8, 9^ Elevated BrainAGE has been associated with cognitive decline and increased risk of neurodegenerative disease, suggesting that it reflects structural vulnerability beyond chronological ageing.^10, 6^ Large-scale studies further indicate that brain ageing is not uniform, but follows multiple neuroanatomical trajectories, including patterns dominated by medial temporal involvement as well as more distributed cortical atrophy affecting frontal, parietal, and occipital regions.^6, 11^ Importantly, such patterns are observed even among cognitively healthy individuals, raising the possibility that early-emerging ageing phenotypes shape later susceptibility to neurodegenerative disease.^12, 13, 3^

Within this framework, the medial temporal lobe (MTL) is of particular interest. Healthy ageing already produces early structural decline in MTL regions such as the hippocampus, entorhinal cortex, and parahippocampal cortex, and these same structures are affected early in Alzheimer’s disease^14, 15, 16^. These observations suggest that regional measures of brain ageing, particularly temporal lobe BrainAGE, may capture early neurobiological changes associated with future cognitive decline. However, it remains unclear whether such regional indices provide additional sensitivity beyond global BrainAGE during the preclinical stages preceding mild cognitive impairment (MCI) or dementia.

Despite substantial research on the development of brain age models and their associations with cognitive decline and neurodegenerative diseases, key questions remain unresolved.^9,17^ It is not yet clear how early increases in BrainAGE can be detected in individuals who are cognitively normal but later decline, nor whether these differences reflect progressive divergence in longitudinal trajectories or stable offsets present years before diagnosis. In addition, most prior work has been conducted in well-controlled research cohorts, leaving open the question of whether similar patterns can be reliably detected in real-world clinical imaging data characterized by heterogeneous acquisition protocols. Finally, the extent to which BrainAGE provides incremental predictive value for progression to MCI or dementia beyond standard demographic, cognitive, and MRI-derived measures has not been fully characterized.

In this study, we address these questions by examining global and temporal lobe BrainAGE in cognitively normal individuals who either remain stable or later progress to MCI or dementia. Using data from the Alzheimer’s Disease Neuroimaging Initiative (ADNI), we apply a validated brain age framework,^11^ analyse longitudinal trajectories of BrainAGE scores, and investigate whether individuals who subsequently convert to MCI or dementia, exhibit early divergence in structural ageing trajectories detectable several years before diagnosis. We further evaluate whether BrainAGE improves prediction of progression to MCI or dementia and future memory change beyond demographic, APOE4, cognitive, and MRI-derived measures. To assess generalizability, key analyses were replicated in OSTPRE (population-based Kuopio Osteoporosis Risk Factor and Prevention Study), with routine clinical MRI data. Together, these analyses evaluate whether BrainAGE can serve as an accessible marker of early divergence in brain ageing trajectories and improve identification of cognitively normal individuals at increased risk of future cognitive decline.

## Materials and Methods

An overview of the study design is shown in Figure 1a. BrainAGE measures were derived from structural MRI and evaluated using four complementary analytical approaches. First, longitudinal analyses were performed to characterize BrainAGE trajectories preceding conversion to MCI or dementia and to examine their association with subsequent changes in memory performance. Second, cross-sectional analyses were performed to compare BrainAGE between groups at predefined time points preceding conversion to MCI or dementia and in relation to subsequent changes in memory performance. Third, time-to-event analyses evaluated whether BrainAGE was associated with the risk of future conversion from cognitively normal status to MCI or dementia. Finally, predictive modelling analyses assessed whether BrainAGE improved prediction of future cognitive outcomes beyond demographic, genetic, cognitive, and MRI-derived measures. Primary analyses were performed in ADNI, and key findings related to progression to MCI or dementia were replicated in the OSTPRE cohort.

**Figure 1.**
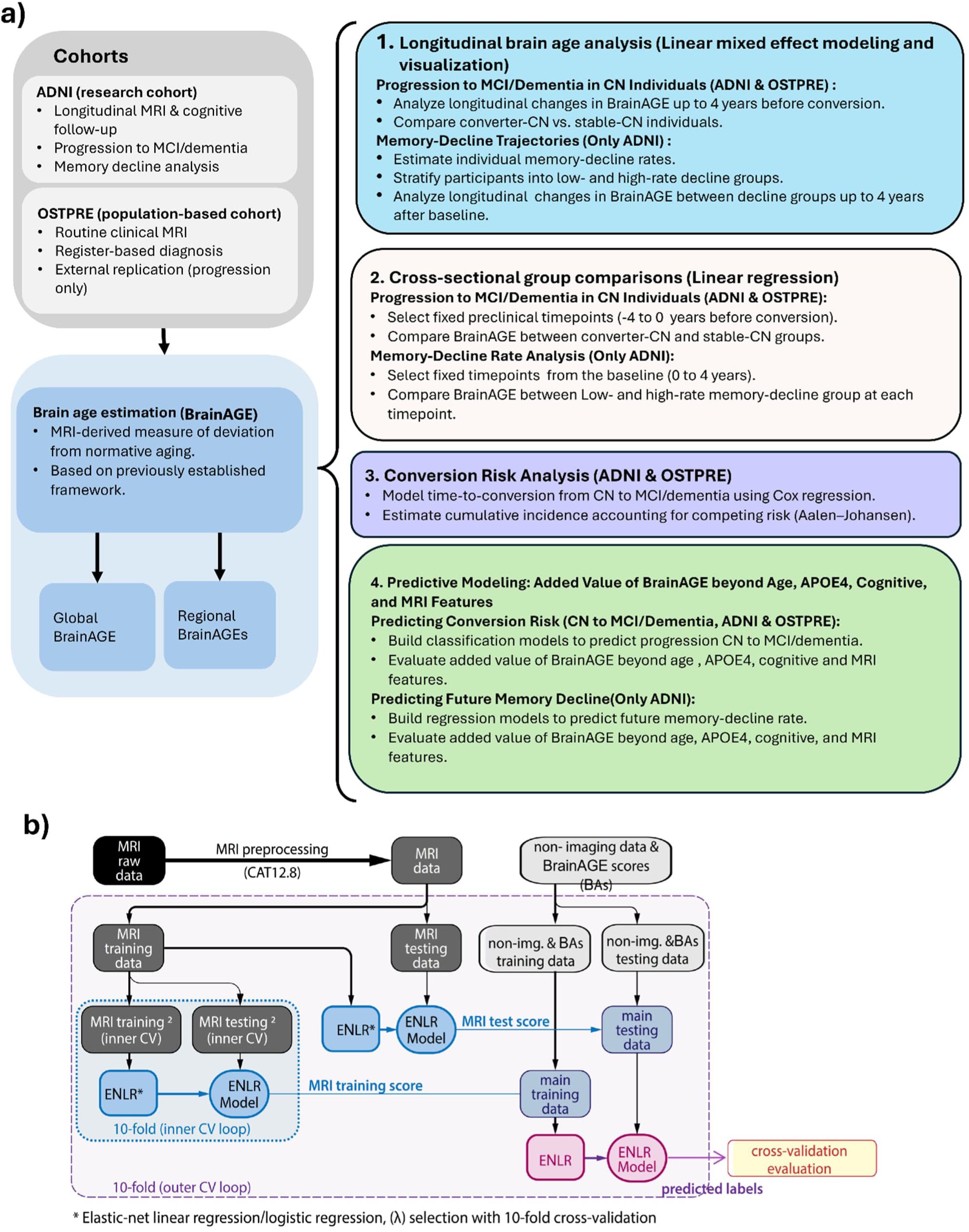
Study workflow. (a) Schematic overview of the study design with the different experimental analyses performed. (b) Schematic representation of the predictive modelling framework, adapted from our previous work.^18^ MCI, mild cognitive impairment; CN, cognitively normal.

### ADNI data

Data for the primary analyses were obtained from the Alzheimer’s Disease Neuroimaging Initiative (ADNI; http://adni.loni.usc.edu). ADNI was launched in 2003 as a publicprivate partnership led by Principal Investigator Michael W. Weiner, MD. The study was designed to determine whether serial magnetic resonance imaging, positron emission tomography, biological markers, and detailed clinical and neuropsychological assessments could be combined to measure the progression of MCI and early AD. Up-to-date information is available at http://www.adni-info.org.

We investigated progression from cognitively normal (CN) status to mild cognitive impairment (MCI) or dementia, as well as longitudinal memory change. For progression analyses, we included participants who were cognitively normal or reported subjective memory complaints at baseline and who had information of APOE4 status, baseline composite cognitive scores, T1-weighted MRI, and longitudinal diagnostic follow-up. Participants were classified as stable CN or converters according to their longitudinal diagnostic trajectory. The final sample included 330 stable individuals and 104 converters. For memory-decline analyses, participants were required to have baseline MRI, APOE4 status, cognitive assessments, and longitudinal memory data. Baseline characteristics are presented in Table 1.

**Table 1.** Baseline characteristics of participants included in the study. Values are mean (standard deviation) unless otherwise indicated.

| A. Conversion analysis (Cognitively normal at baseline) |  |  |  |  |
| --- | --- | --- | --- | --- |
| Characteristic | ADNI |  | OSTPRE |  |
|  | Stable-CN | Converter-CN | Stable-CN | Converter-CN |
| Sample size, n | 330 | 104 | 521 | 387 |
| Age at baseline, years | 71.50 (6.11) | 75.35 (5.10) | 74.37 (4.09) | 75.74 (4.40) |
| Sex, M/F | 137/193 | 58/46 | All women | All women |
| APOE4 (0/1/2) | 237/86/7 | 70/32/2 | – | – |
| Global BrainAGE, years | -1.13 (3.20) | 0.86 (3.91) | -2.22 (4.24) | 0.23 (4.00) |
| Temporal BrainAGE, years | -1.42 (3.28) | 0.51 (3.79) | -2.83 (4.88) | 0.22 (5.24) |
| Composite memory score (MEM) | 0.98 (0.43) | 0.60 (0.41) | – | – |
| Composite language score (LAN) | 0.94 (0.54) | 0.60 (0.47) | – | – |
| Composite executive function score (EXF) | 0.96 (0.51) | 0.65 (0.48) | – | – |
| Cognitive status (CN/SMC) | 205/125 | 79/25 | 397/124 | 231/156 |
| B. Memory decline analysis (Participants without dementia) |  |  |  |  |
| Characteristic | ADNI |  |  |  |
|  | CN | MCI |  |  |
| Sample size, n | 655 | 783 |  |  |
| Age at baseline, years | 73.03 (6.19) | 72.79 (7.48) |  |  |
| Sex, M/F | 294/361 | 468/315 |  |  |
| APOE4 (0/1/2) | 453/185/17 | 406/295/82 |  |  |
| Global BrainAGE, years | -0.35 (3.36) | 1.90 (3.74) |  |  |
| Temporal BrainAGE, years | -0.66 (3.44) | 1.99 (4.05) |  |  |
| Composite memory score (MEM) | 0.87 (0.46) | 0.13 (0.51) |  |  |
| Composite language score (LAN) | 0.83 (0.55) | 0.39 (0.52) |  |  |
| Composite executive function score (EXF) | 0.84 (0.52) | 0.40 (0.58) |  |  |

CN, cognitively normal; SMC, subjective memory complaints; MCI, mild cognitive impairment.

Detailed participant selection criteria, diagnostic definitions, longitudinal follow-up procedures, construction of the memory-decline cohort, and data sources are provided in the Supplementary Methods (Section S1.1). The details of MRI acquisition are reviewed in.^19^

### OSTPRE

The Kuopio Osteoporosis Risk Factor and Prevention Study (OSTPRE) is a population-based cohort of women living in Eastern Finland.^20–24^ The study has been approved by the Ethics Committee of Kuopio University Hospital. Permissions for register data have been obtained from the Finnish data permit authority for social and health care data (Dnro THL/6840/14.02.00/2020). The study was performed in accordance with the ethical standards of the 1964 Declaration of Helsinki and its later amendments.

Brain MRI data for OSTPRE women were obtained from routine clinical imaging archived in the regional picture archiving and communication system (PACS), and cognitive status was determined through linkage with national health registers. Brain MRI data in the OSTPRE cohort was obtained from routine clinical imaging archived in the regional picture archiving and communication system (PACS), and cognitive status was determined through linkage with national health registers. Analyses were based on whole-brain T1-weighted structural MRI scans that passed automated quality control. Images were acquired on both 1.5 T and 3.0 T scanners from multiple vendors, predominantly Siemens, followed by Philips and Toshiba. Further details of MRI acquisition and quality control have been described previously.^25^

OSTPRE was used as an external replication cohort for progression to MCI or dementia analyses only. Memory-change analyses could not be replicated in OSTPRE because harmonised longitudinal cognitive composite measures were unavailable. For progression to MCI or dementia analyses, participants were required to be cognitively normal at their first available MRI examination, defined as having no memory complaints or subjective memory complaints, and were classified as stable or converters according to subsequent register-based diagnoses of MCI or dementia. The final sample included 521 stable individuals and 387 converters (Table 1). Further details regarding cohort construction, diagnostic ascertainment, follow-up definitions, scan-selection procedures, and the handling of variable follow-up durations are provided in the Supplementary Methods (Section S1.2).

### MRI preprocessing and BrainAGE model

Structural T1-weighted brain MRI scans were preprocessed using the Computational Anatomy Toolbox (CAT12, version 8.1; https://neuro-jena.github.io/cat/) implemented in SPM (https://www.fil.ion.ucl.ac.uk/spm/), following the framework described by.^18^ Default CAT12 preprocessing settings were applied. Images were segmented into grey matter (GM) and white matter (WM) and non-linearly normalised to stereotactic space using the shooting approach.^26^ Based on the spatially normalised GM and WM segments, regional volumetric measures were extracted using the Neuromorphometrics atlas, derived from the MICCAI 2012 Multi-Atlas Labeling Challenge dataset. In total, 136 regional volumetric features were obtained. Intracranial volume (ICV) was included as an additional feature rather than used solely for normalization.^27^

Cortical thickness was estimated as described by Dahnke et al.^28^ and registered to the FSaverage surface template.^29^ Regional cortical thickness measures were then derived using the Desikan– Killiany (DK40) atlas,^30^ resulting in 69 regionally averaged features.

BrainAGE was estimated using a previously validated BrainAGE framework based on Gaussian process regression.^11^ The model was trained on healthy adult participants from five publicly available datasets (IXI (https://brain-development.org/ixi-dataset/), OASIS-3,^31^ NKI Enhanced (https://fcon1000.projects.nitrc.org/indi/pro/eNKI_RS_TRT/FrontPage.html), SALD,^32^ and Cam-CAN^33^) to predict chronological age from structural brain features. For OSTPRE, scanner effects were corrected during BrainAGE estimation to account for the heterogeneous clinical MRI data, as described previously.^11, 24^ No scanner correction was applied to ADNI because MRI acquisition followed a highly standardised multicentre protocol. Predicted brain age values were corrected for age-related bias using a linear adjustment as described by.^34^ BrainAGE was defined as the difference between the bias-corrected predicted brain age and chronological age, with positive values indicating an older-appearing brain.

In addition to global BrainAGE, regional BrainAGE estimates were computed using the same modelling framework, restricted to features from predefined anatomical regions. Specifically, GM segments were partitioned into 10 lobes (5 per hemisphere) based on the Brain lobe atlas,^35^ and separate Gaussian process regression models were trained for each region. This approach provides spatially specific estimates of deviations in brain ageing. The temporal lobe BrainAGE estimate (hereafter referred to as Temporal BrainAGE) was used as the primary regional BrainAGE measure in this study.

### Statistical analysis

Longitudinal changes in global and regional BrainAGE were examined using linear mixed-effects models.^36^ BrainAGE was modelled as the dependent variable, with fixed effects for time, group (converter-CN versus stable-CN or high-rate versus lowrate memory decliners), and their interaction. A four-year observation window was used for all longitudinal analyses. For progression analyses, trajectories were aligned relative to the time of conversion to MCI or dementia, whereas memory-change analyses were aligned relative to baseline. The interaction term was used to assess whether BrainAGE trajectories differed between groups over time. In ADNI, models were adjusted for APOE4 status and sex, whereas such adjustments were not possible in OSTPRE because APOE4 data were unavailable and all participants were women. Participants were required to have at least two MRI scans within the relevant analysis window. Further details regarding time alignment, participant inclusion, and model specification are provided in the Supplementary Methods.

To characterise BrainAGE differences at predefined time points before conversion to MCI or dementia and during follow-up for memory change, we performed cross-sectional analyses at predefined time points using multivariable linear regression models. For progression analyses, comparisons were performed at annual intervals during the four years preceding conversion, whereas memory-change analyses examined annual intervals during the four years following baseline. Discriminative performance at each time point was assessed using receiver operating characteristic (ROC) analysis, with BrainAGE as the predictor and group status as the outcome and summarized using the area under the curve (AUC).

Associations between BrainAGE and progression from cognitively normal status to MCI or dementia were examined using cause-specific Cox proportional hazards models. BrainAGE was analysed separately for Global BrainAGE and Temporal BrainAGEs, and hazard ratios (HRs) with 95% confidence intervals (CIs) were reported. To account for death as a competing event, cumulative incidence functions were estimated using competing-risk methodology and compared between groups.

To evaluate the robustness of the findings, sensitivity analyses were performed in a restricted subset of participants with BrainAGE measurements available at consistent follow-up time points. Detailed descriptions of model specification, time-alignment procedures, competing-risk analyses, sensitivity analyses, and statistical implementation are provided in the Supplementary Methods (Section S1.3).

### Integrating BrainAGE with demographic, APOE4, cognitive, and MRI predictors

To evaluate whether BrainAGE provided incremental predictive value beyond demographic, APOE4, cognitive, and MRI-derived features, predictive analyses were performed for (i) progression from CN status to MCI or dementia and (ii) annual change in memory performance (MEM composite score) among ADNI participants without dementia. All models used baseline predictors only.

Participants were included if they had a baseline T1-weighted MRI, BrainAGE estimates, and all required non-imaging predictors. In ADNI, these included age, APOE4 status, and baseline composite cognitive scores. In OSTPRE, APOE4 and composite cognitive measures were unavailable; therefore, baseline cognitive status (no memory complaints versus subjective memory complaints) was used as a proxy for cognitive variation. Outcomes were determined from longitudinal follow-up.

A two-stage machine-learning framework was implemented following Moradi et al.^18^ to integrate high-dimensional MRI data with lower-dimensional non-imaging predictors. Because structural MRI measures (regional volumes and cortical thickness) were substantially higher-dimensional than other predictors, baseline MRI features were first reduced to a task-specific MRI-derived score using ENLR. This MRI-derived score was then combined with BrainAGE measures and non-imaging predictors to evaluate their joint predictive contributions. For regional BrainAGE analyses, left and right hemisphere values were averaged because of the high inter-hemispheric correlation (Supplementary Figure S2). Although both BrainAGE and the MRI-derived score were derived from structural MRI, they reflected different modelling objectives: BrainAGE quantified deviation from normal ageing, whereas the MRI-derived score was optimised for prediction of the outcome of interest.

To evaluate the incremental contribution of BrainAGE, four predefined models with increasing information content were compared. In ADNI, the Basic model included age, APOE4 status, and baseline composite cognitive scores. The MRI model extended the Basic model by adding the MRI-derived score. The GlobalBA model further added Global BrainAGE, and the RegionalBA model additionally included regional BrainAGE measures. Because composite cognitive scores and APOE4 were not available in OSTPRE, the Basic model used in ADNI could not be replicated. Instead, an MRI-based model was defined that included age, baseline cognitive status, and MRI-derived score with Global BrainAGE and regional BrainAGE added sequentially in the GlobalBA and RegionalBA models.

Task-specific elastic net models were then fitted for each outcome, and model performance was evaluated using repeated nested cross-validation. Classification performance was assessed using the area under the receiver operating characteristic curve (AUC), whereas prediction of annual memory change was evaluated using Pearson correlation and mean absolute error (MAE). This framework is illustrated in Figure 1b. Additional details regarding predictor construction, model implementation, performance comparisons, and statistical testing are provided in the Supplementary Methods (Section S1.4).

## Results

As outlined in Figure 1a, we first characterised BrainAGE using complementary longitudinal, cross-sectional, and time-to-event analyses to examine its relationship with progression to MCI or dementia and memory change. We then evaluated the predictive value of BrainAGE beyond demographic, genetic, cognitive, and MRI-derived measures.

### BrainAGE diverges years before MCI/dementia diagnosis

In ADNI, longitudinal mixed-effects models adjusted for APOE4 status and sex showed that both Global and Temporal BrainAGE increased over the four years preceding conversion to MCI or dementia. Global BrainAGE increased by *β* = 0.11 units per year (95% CI 0.07 to 0.15; *t* = 5.55, *p <* 0.001), whereas Temporal BrainAGE increased more steeply (*β* = 0.21 units per year, 95% CI 0.17 to 0.24; *t* = 10.80, *p <* 0.001). Individuals who subsequently converted had higher BrainAGE at the time of conversion than stable cognitively normal participants (Global: *β* = 3.29, 95% CI 2.43 to 4.15; *t* = 7.45, *p <* 0.001; Temporal: *β* = 3.37, 95% CI 2.48 to 4.26; *t* = 7.39, *p <* 0.001). Converters also showed steeper longitudinal increases in BrainAGE, reflected by significant group by time interactions (Global: *β* = 0.17, 95% CI 0.07 to 0.26; *t* = 3.49, *p <* 0.001; Temporal: *β* = 0.23, 95% CI 0.14 to 0.33; *t* = 5.01, *p <* 0.001). Detailed results are provided in Supplementary Table S2.

Cross-sectional analyses at predefined time points preceding conversion to MCI or dementia showed higher BrainAGE in converters at all examined time points. At −4 years, the adjusted group difference was *β* = 2.83 for Global BrainAGE (95% CI 1.80 to 3.87) and *β* = 3.09 for Temporal BrainAGE (95% CI 2.05 to 4.13), with similar effect sizes at later time points (Supplementary Table S3). Using BrainAGE alone, cross-sectional discrimination yielded AUC values ranging from 0.69 to 0.75 for Global BrainAGE and from 0.65 to 0.73 for Temporal BrainAGE. At −4 years, the AUC was 0.72 (95% CI 0.63 to 0.81) for Global BrainAGE and 0.73 (95% CI 0.66 to 0.81) for Temporal BrainAGE. Detailed results are provided in Supplementary Table S3.

Sensitivity analyses in a restricted ADNI subset with BrainAGE measurements available for all participants at −4, −2, and 0 years (15 converters and 117 stable individuals) showed similar findings. Global BrainAGE increased by *β* = 0.10 units per year (95% CI 0.04 to 0.16; *t* = 3.49, *p <* 0.001), and converters had higher BrainAGE at conversion (*β* = 4.30, 95% CI 2.41 to 6.19; *t* = 4.50, *p <* 0.001). Significant group-by-time interactions remained for both Global BrainAGE (*β* = 0.30 per year, 95% CI 0.13 to 0.47; *t* = 3.47, *p <* 0.001) and Temporal BrainAGE (*β* = 0.32 per year, 95% CI 0.16 to 0.48; *t* = 4.03, *p <* 0.001), indicating that converters continued to have accelerated brain ageing relative to stable individuals (Supplementary Table S4). Cross-sectional analyses at predefined time points showed comparable discriminative performance up to four years before conversion. At −4 years, the AUC was 0.74 (95% CI 0.61 to 0.86) for Global BrainAGE and 0.68 (95% CI 0.55 to 0.81) for Temporal BrainAGE (Supplementary Table S5). These findings confirmed the robustness of the primary analyses in this strictly matched longitudinal sample.

Figure 2 illustrates the longitudinal Global BrainAGE patterns in ADNI and OSTPRE. Raincloud plots showed higher Global BrainAGE values in converters than in stable participants across all examined time points. Because participants did not contribute MRI scans at every time point, longitudinal slope differences were less directly apparent in the full-sample visualisation. Raincloud plots restricted to individuals with complete longitudinal MRI data (Supplementary Figure S3) more clearly illustrated the steeper BrainAGE trajectories in converters. Similar patterns were observed for Temporal BrainAGE (Supplementary Figure S4).

**Figure 2.**
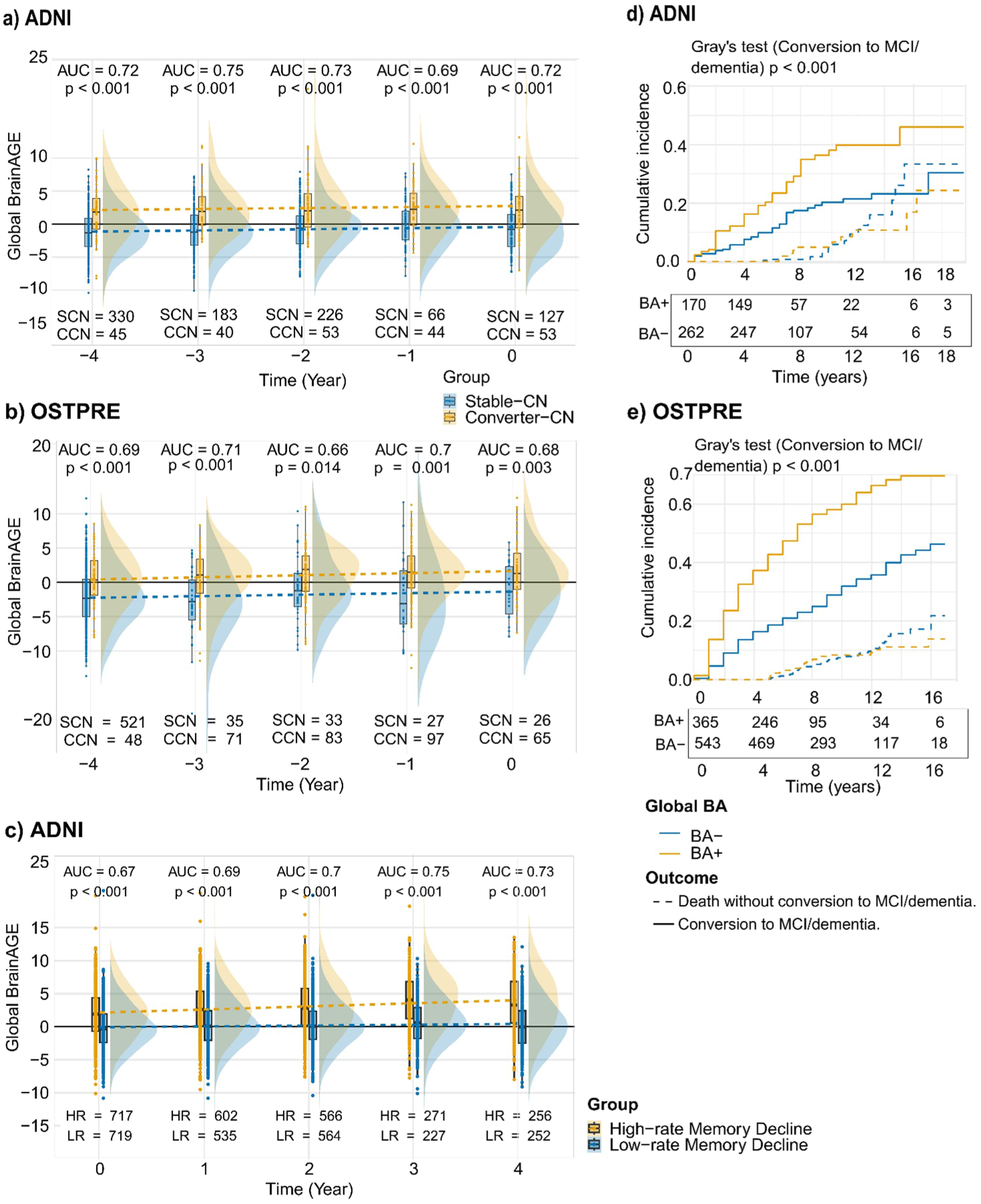
Longitudinal trajectories of Global BrainAGE and risk of conversion in ADNI and OSTPRE. (a,b) Global BrainAGE values aligned relative to conversion to MCI or dementia in cognitively normal individuals who remained stable (stable-CN) or subsequently converted (converter-CN) in ADNI (a) and OSTPRE (b), shown as raincloud plots with fitted group trajectories. Sample sizes and cross-sectional AUC values are shown at each time point. (c) Global BrainAGE values in ADNI among individuals without dementia, stratified according to subsequent change in memory performance. (d,e) Cumulative incidence of conversion to MCI or dementia estimated using the Aalen-Johansen estimator (accounting for death as a competing event) in ADNI (d) and OSTPRE (e), stratified by baseline Global BrainAGE (BA+ and BA-). Numbers at risk are shown below each panel. BA+ denotes Global BrainAGE > 0.

### BrainAGE divergence replicates in a population-based cohort

Longitudinal BrainAGE estimates in OSTPRE were based on fewer repeated MRI measurements per participant than in ADNI because of differences in data density and follow-up structure between the cohorts (see Methods). In OSTPRE, longitudinal mixed-effects models showed increasing BrainAGE over time. Temporal BrainAGE increased significantly (*β* = 0.33 per year, 95% CI 0.05 to 0.62, *t* = 2.27, *p* = 0.024), whereas the increase in Global BrainAGE was not statistically significant (*β* = 0.17 per year, 95% CI −0.11 to 0.45, *t* = 1.18, *p* = 0.241). Individuals who subsequently converted to MCI or dementia had higher BrainAGE at the time of conversion (Global: *β* = 3.76, 95% CI 1.84 to 5.69, *t* = 3.83, *p* = 0.0002; Temporal: *β* = 3.96, 95% CI 1.99 to 5.93, *t* = 3.92, *p* = 0.0001). However, there was no statistically significant evidence of a differential rate of change between groups for either Global BrainAGE (*β* = 0.37 per year, 95% CI −0.12 to 0.86, *t* = 1.50, *p* = 0.134) or Temporal BrainAGE (*β* = 0.16 per year, 95% CI −0.34 to 0.66, *t* = 0.62, *p* = 0.537) (Supplementary Table S6).

Cross-sectional analyses confirmed elevated BrainAGE in converters up to four years before conversion to MCI or dementia (Supplementary Table S7). Group differences were consistently observed for both Global and Temporal BrainAGE across all time points (all *p <* 0.05), with larger effect sizes for Temporal BrainAGE. Discriminative performance was moderate for Global BrainAGE (AUC range 0.66–0.71) and higher for Temporal BrainAGE (AUC range 0.69–0.79).

Consistent with these findings, raincloud plots (Figure 2b, and Supplementary Figure S4) show higher BrainAGE values in converters compared to stable individuals across all examined time points for both Global and Temporal measures.

### Elevated BrainAGE is associated with increased risk of conversion to MCI or dementia

Across both ADNI and OSTPRE cohorts, BrainAGE positivity was consistently associated with an increased risk of conversion from CN status to MCI or dementia. Cause-specific Cox proportional hazards analyses showed that BrainAGE positivity was associated with an increased risk of conversion in both cohorts. In ADNI, Global BrainAGE positivity was associated with a hazard ratio (HR) of 1.98 (95% CI 1.34 to 2.91, *p <* 0.001), and Temporal BrainAGE with an HR of 1.60 (95% CI 1.09 to 2.36, *p* = 0.018). In OSTPRE, Global BrainAGE positivity yielded an HR of 2.58 (95% CI 2.11 to 3.16) and Temporal BrainAGE an HR of 3.05 (95% CI 2.55 to 3.73), both *p <* 0.0001. The proportional hazards assumption was evaluated using Schoenfeld residuals. No violations were observed for Global or Temporal BrainAGE in ADNI or for Global BrainAGE in OSTPRE. A deviation from the proportional hazards assumption was observed for Temporal BrainAGE in OSTPRE (p = 0.001), indicating potential time-dependent effects. Visual assessment is provided in Supplementary Fig S5.

These findings were supported by cumulative incidence analyses. Aalen–Johansen cumulative incidence curves (Figure 2d,e) demonstrated clear separation between BrainAGEpositive and BrainAGE-negative individuals for Global BrainAGE in both cohorts (Gray’s test *p <* 0.001), indicating a higher cumulative incidence of conversion among individuals with elevated BrainAGE while accounting for death as a competing risk. Similar patterns were observed for Temporal BrainAGE, with corresponding cumulative incidence curves shown in Supplementary Figure S6. Supplementary Table S8 summarizes hazard ratios for all models.

Comparing the associations of Global and Temporal BrainAGE with conversion, in ADNI, Global BrainAGE showed greater separation between groups compared to Temporal BrainAGE, consistent with its higher hazard ratio. In contrast, in OSTPRE, Temporal BrainAGE showed greater separation and was associated with the highest hazard ratio for conversion, albeit with some evidence of time-varying risk.

### Accelerated structural brain ageing tracks cognitive decline

Among non-demented participants in ADNI, linear mixed-effects models adjusted for APOE4 and sex showed that individuals with faster memory decline had higher BrainAGE and steeper increases over time. Group differences were *β* = 2.04 for Global BrainAGE (95% CI 1.64 to 2.44; t = 10.03, p < 0.001) and *β* = 2.54 for Temporal BrainAGE (95% CI 2.12 to 2.97; t = 11.77, p < 0.001). Significant group-by-time interactions indicated faster longitudinal increases in individuals with faster memory decline (Global: *β* = 0.31 per year, 95% CI 0.27 to 0.35; t = 17.08, p < 0.001; Temporal: *β* = 0.37 per year, 95% CI 0.33 to 0.40; t = 20.60, p < 0.001) (Supplementary Table S9).

Cross-sectional analyses showed increasing group differences from baseline to year four. For Global BrainAGE, the difference increased from *β* = 2.11 at baseline to *β* = 3.24 at year four. For Temporal BrainAGE, the difference increased from *β* = 2.56 to *β* = 3.97. Correspondingly, AUC values increased from 0.67 to 0.73 for Global BrainAGE and from 0.69 to 0.76 for Temporal BrainAGE (Supplementary Table S10).

Sensitivity analyses restricted to individuals with complete data at 0, 2, and 4 years (*n* = 484) yielded consistent findings. Individuals with faster memory decline had higher baseline BrainAGE (Global: *β* = 2.27, 95% CI 1.62 to 2.91; Temporal: *β* = 2.73, 95% CI 2.03 to 3.43; both *p <* 0.001) and steeper increases over time (Global: *β* = 0.29 per year; Temporal: *β* = 0.35 per year; both *p <* 0.001) (Supplementary Table S11). Cross-sectional group differences and AUC values showed similar patterns, with larger differences for Temporal BrainAGE (Supplementary Table S12).

Figure 2c illustrated Global BrainAGE trajectories in individuals with high and low memory decline, showing consistently higher BrainAGE values and steeper increases over time in the high-decline group. Corresponding raincloud plots for Temporal BrainAGE, together with plots from the subset with complete four-year longitudinal MRI data, are shown in Supplementary Figure S7.

### BrainAGE improves prediction of future cognitive outcomes

We next evaluated whether BrainAGE improved prediction of future cognitive outcomes beyond demographic, APOE4, cognitive, and MRI-derived measures. Four predefined predictive models with increasing information content (Basic, MRI, GlobalBA, and RegionalBA; see Methods) were compared using repeated nested cross-validation.

#### Prediction of progression to MCI or dementia

In ADNI, the Basic model achieved an AUC of 0.75 (95% CI 0.70 to 0.80). Adding MRI modestly improved performance (AUC 0.78, 95% CI 0.73 to 0.83). Incorporating Global BrainAGE further increased performance (AUC 0.80, 95% CI 0.75 to 0.84), whereas the RegionalBA model performed similarly (AUC 0.79, 95% CI 0.73 to 0.83). DeLong tests on the median-performing cross-validation run showed that the GlobalBA model outperformed both the Basic (*p* = 0.03) and MRI (*p* = 0.02) models, whereas no difference was observed between the MRI and RegionalBA models (*p* = 0.82). Additional performance metrics are reported in Supplementary Figure S8.

To examine predictions across different time horizons, converters were stratified according to time to conversion. For short-term converters (conversion within four years), all models performed similarly (AUCs 0.85–0.88; all *p >* 0.05). For long-term converters (conversion more than four years after baseline), performance was lower for the Basic model (AUC 0.67) and improved after adding MRI (AUC 0.71, *p* = 0.038 versus Basic) and Global BrainAGE (AUC 0.73, *p* = 0.008 versus Basic). The RegionalBA model achieved an AUC of 0.72 (Figure 3a). The ADNI cohort included 48 short-term and 56 long-term converters, with time to conversion extending up to 17 years after baseline (Figure 3b).

**Figure 3.**
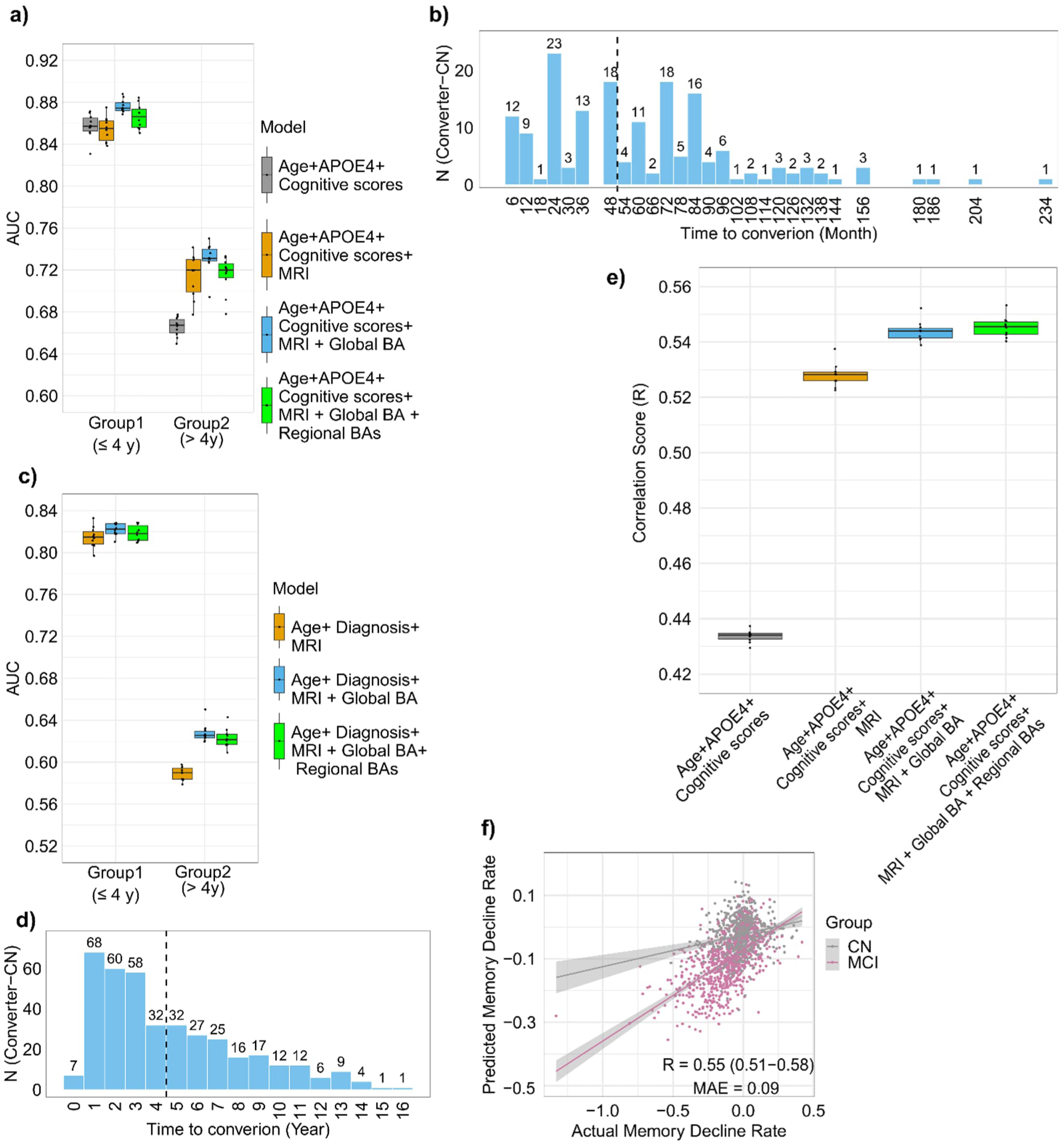
Predictive performance of the prediction models in ADNI and OSTPRE. (a, c) Distributions of AUC values for short-term and long-term prediction of progression from cognitively normal status to MCI or dementia across the evaluated models in ADNI (a) and OSTPRE (c). (b, d) Distribution of converters according to time from baseline to conversion in ADNI (b; months) and OSTPRE (d; years). (e) Distribution of Pearson correlation coefficients ($R$) between predicted and observed annual memory change across repeated nested cross-validation runs in ADNI. (f) Predicted versus observed annual memory change for the RegionalBA model from the median-performing cross-validation run in ADNI.

In OSTPRE, APOE4 status and composite cognitive scores were unavailable; therefore, the Basic model could not be replicated. The MRI model achieved an AUC of 0.70 (95% CI 0.67 to 0.73). Adding Global BrainAGE increased performance to 0.72 (95% CI 0.69 to 0.76), showing a statistical trend (*p* = 0.056). Similar performance was observed for the RegionalBA model (AUC 0.72, 95% CI 0.69 to 0.75). For short-term converters, AUC values ranged from 0.81 to 0.82, whereas long-term prediction was more challenging (AUC 0.56–0.60). Nevertheless, inclusion of BrainAGE numerically improved long-term prediction (*p* = 0.048) (Figure 3c). The OSTPRE cohort included 225 short-term and 162 long-term converters, with time to conversion extending up to 16 years after baseline (Figure 3d).

#### Prediction of annual memory change

Prediction of annual memory change showed a similar pattern. The Basic model achieved a correlation between predicted and observed memory change of *R* = 0.43 (95% CI 0.39 to 0.48) with a mean absolute error (MAE) of 0.10 (95% CI 0.095 to 0.105). Adding the MRI-derived score improved performance (*R* = 0.53, 95% CI 0.49 to 0.56; MAE = 0.094, 95% CI 0.089 to 0.098). Incorporating Global BrainAGE further improved prediction (*R* = 0.54, 95% CI 0.51 to 0.58; MAE = 0.093, 95% CI 0.088 to 0.097), whereas the RegionalBA model achieved the highest correlation (*R* = 0.55, 95% CI 0.51 to 0.58) with a similar MAE (0.093, 95% CI 0.088 to 0.097). Improvements from the Basic model to the MRI, GlobalBA, and RegionalBA models were all statistically significant (*p <* 0.001), and both BrainAGE models outperformed the MRI model (*p <* 0.01), whereas no difference was observed between the GlobalBA and RegionalBA models.

Figure 3e shows the distribution of correlation coefficients across repeated nested cross-validation runs. Figure 3f shows observed versus predicted annual memory change for the RegionalBA model. Stronger associations were observed in individuals with MCI whereas the smaller range of cognitive change among cognitively normal participants made prediction more challenging.

### Relative contribution of cognitive, MRI, and BrainAGE predictors

To assess the relative importance of individual predictors, we examined the mean absolute elastic-net regression coefficients averaged across cross-validation folds and repetitions for individual predictors (Figure 4). In the ADNI conversion prediction, the largest coefficients were observed for the memory composite and the MRI-derived score, followed by language composites and executive function. Among the BrainAGE predictors, Frontal, Temporal, and Global BrainAGE contributed most, followed by Occipital BrainAGE, whereas Parietal and Subcortical/Cerebellar BrainAGE showed smaller contributions. Age and APOE4 had relatively modest coefficients (Figure 4a).

**Figure 4.**
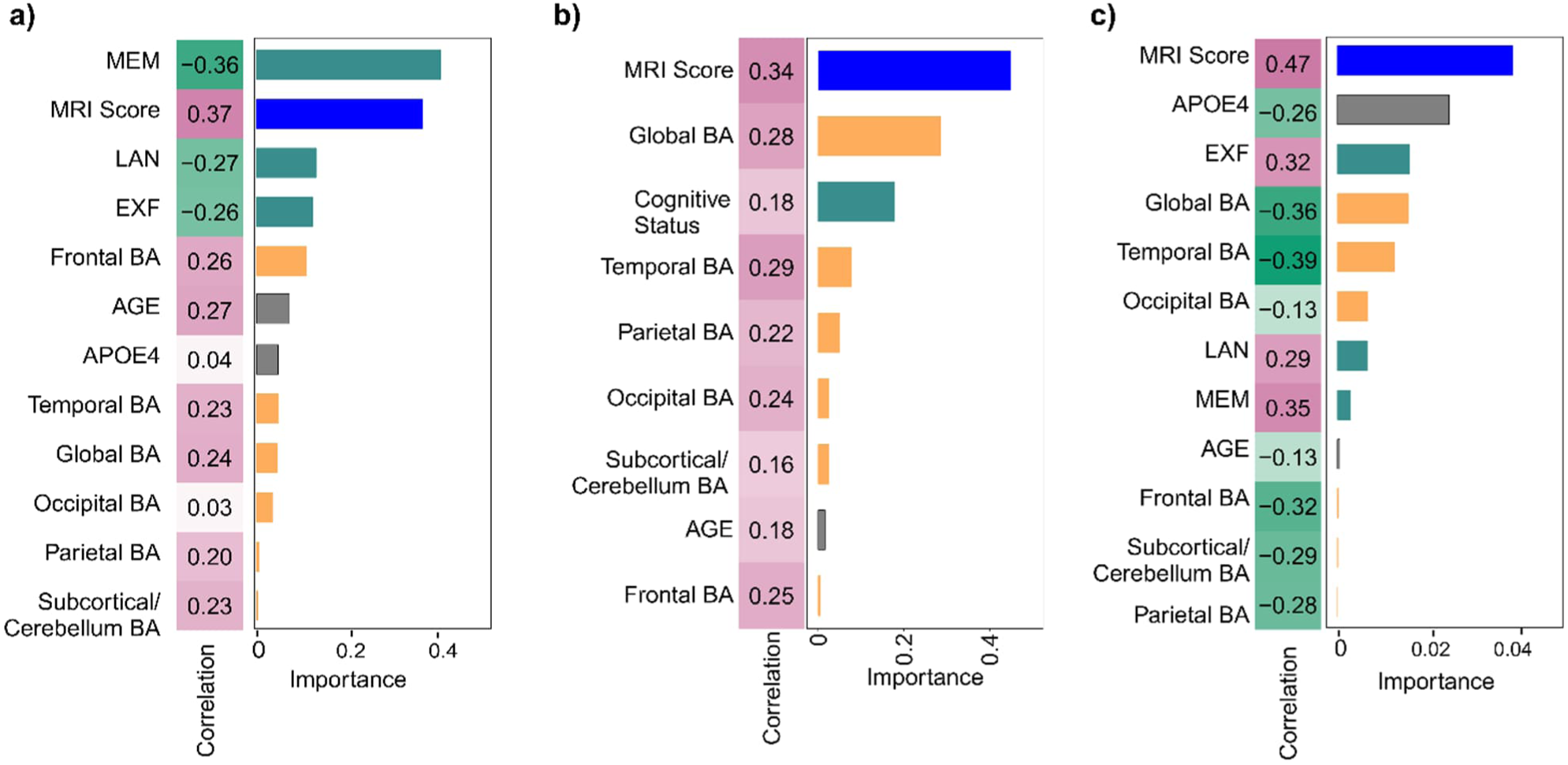
Predictor importance in regional BrainAGE models for conversion and memory outcomes. Predictor importance was estimated as the mean absolute elastic-net regression coefficients averaged across 10 repetitions of 10-fold cross-validation. Pearson correlation coefficients between each predictor and the corresponding outcome are also shown. The MRI-derived score represents the cross-validated prediction score obtained by applying elastic-net regression to high-dimensional MRI measures. (a) Regional BrainAGE model for predicting progression from cognitively normal status to MCI or dementia in ADNI; (b) corresponding analysis in OSTPRE; (c) Regional BrainAGE model for predicting annual memory change in non-demented individuals in ADNI. MEM, composite memory score; LAN, composite language score; EXF, composite executive function score.

In OSTPRE, where composite cognitive scores and APOE4 were unavailable, the MRI-derived score and global BrainAGE were most influential, followed by baseline cognitive status and Temporal BrainAGE. Age showed smaller coefficients, whereas Frontal BrainAGE had the smallest contribution (Figure 4b).

For prediction of annual memory change in ADNI, the MRI-derived score had the largest coefficient, followed by APOE4 status and baseline executive function. Temporal and Global BrainAGE contributed more strongly than in the conversion prediction models and had contributions comparable to baseline executive function. The language and memory composites together with Occipital BrainAGE showed moderate contributions, whereas age, Frontal BrainAGE, Parietal BrainAGE, and Subcortical/Cerebellar BrainAGE contributed least (Figure 4c).

## Discussion

In this study, we showed that elevated BrainAGE is detectable several years before conversion to MCI or dementia, while individuals are still cognitively normal. Across two complementary cohorts, a research cohort (ADNI) and a population-based cohort (OSTPRE), individuals who later progressed to MCI or dementia had higher BrainAGE values, and in ADNI also showed steeper increases over time. BrainAGE was also associated with subsequent memory decline, with individuals showing faster memory decline having higher BrainAGE values and steeper longitudinal increases. These findings indicate that divergence from normal brain ageing trajectories begins before the onset of clinical cognitive impairment.

An important question arising from these findings is whether these early structural differences are useful for predicting future cognitive outcomes beyond conventional MRI measures. To address this, we evaluated whether BrainAGE improved prediction beyond demographic, APOE4, cognitive, and MRI-derived volumetric and thickness measures. Although the improvements were modest, adding Global BrainAGE consistently improved prediction of both progression to MCI or dementia and annual memory change. This is notable because BrainAGE itself is derived from structural MRI, indicating that it captures information that is not fully represented by conventional regional volumetric and cortical thickness measures. Rather than replacing standard MRI features, BrainAGE provides a complementary summary of distributed structural deviations from normal brain ageing.

The predictive value of BrainAGE depended on the prediction horizon. The greatest improvement was observed for individuals who converted after longer follow-up, a particularly challenging setting in which models based on baseline demographic, genetic, cognitive, and MRI-derived measures typically achieve lower predictive accuracy, as reported in previous studies.^37–39^ In contrast, BrainAGE provided little additional benefit for short-term converters, for whom these baseline measures already showed relatively strong predictive performance. Consequently, the overall improvement in predictive performance for progression to MCI or dementia was modest. This is expected given the inherent difficulty of predicting future cognitive decline from a cognitively normal baseline, particularly over longer prediction horizons, as reported in previous studies.^40, 41^ These findings suggest that BrainAGE captures structural information beyond that provided by conventional MRI measures. Structural MRI contains a large number of regional volumetric and cortical thickness measures, many of which are highly correlated and individually capture only part of the ageing process. BrainAGE summarises these distributed structural patterns as a single measure of deviation from normal brain ageing, providing a biologically meaningful summary that complements conventional MRI features.

Our findings are consistent with previous studies linking elevated brain age to cognitive decline and neurodegeneration, but extend this work in several important ways. Previous studies have largely relied on cross-sectional comparisons or compared cognitively healthy individuals with symptomatic groups.^2,6,10,42^ Here, we demonstrated that BrainAGE differences are already detectable years before the diagnosis of MCI or dementia, while individuals remain cognitively normal, and that these differences continue to evolve over time. At the same time, the observed effects were modest. Considerable overlap remained between groups, and predictive performance was moderate. This is consistent with the multifactorial nature of brain ageing, which is influenced by vascular health, lifestyle, genetics, comorbidities, and other factors that contribute to substantial inter-individual variability^43^.

We also examined regional BrainAGE, focusing on the temporal lobe because of its well-established vulnerability in ageing and Alzheimer’s disease. Temporal BrainAGE showed patterns similar to Global BrainAGE and, in some analyses, slightly stronger associations with cognitive decline. However, regional BrainAGE did not consistently improve prediction beyond Global BrainAGE. This likely reflects the substantial overlap in information captured by global and regional BrainAGE measures, which were highly correlated. Global and regional BrainAGE therefore appear to capture related aspects of structural vulnerability, with regional measures reflecting more localised changes and Global BrainAGE summarising broader patterns of brain ageing^44–47^.

Replication of the main findings in the population-based OSTPRE cohort supports the generalisability of BrainAGE beyond highly controlled research settings. Crosssectional differences between stable individuals and converters were consistent with those observed in ADNI, whereas longitudinal trajectory differences were less pronounced. This likely reflects differences in study design, including fewer repeated MRI examinations, greater variability in scan timing, and increased scanner heterogeneity. These factors reduce sensitivity for detecting longitudinal change but also better represent routine clinical imaging, increasing the clinical relevance of the findings. Variability in follow-up duration and less standardised assessment schedules may also influence the classification of stable individuals and converters in observational cohorts such as OSTPRE. Some individuals classified as stable may convert after the observation period, whereas others may have experienced cognitive decline before receiving a formal diagnosis. These factors are difficult to avoid in longitudinal observational studies and would be expected to attenuate group differences.

Importantly, the associations observed here should not be interpreted as causal. BrainAGE reflects deviations from normative structural ageing but does not identify the biological mechanisms responsible for these differences. It likely captures a combination of ageing-related and disease-related processes and should therefore be viewed as a marker of early structural vulnerability rather than a direct measure of neurodegeneration.

Several limitations should be considered. Although ADNI provides rich phenotyping and structured longitudinal follow-up, its participants may not fully represent the general population. In contrast, OSTPRE reflects a real-world population but includes only women and lacks harmonised cognitive measures and APOE4 status, limiting direct comparisons between cohorts. Differences in data structure and covariate availability may therefore contribute to between-cohort differences in the observed effects. In addition, because conversion occurred at different times across individuals, longitudinal analyses focused on the four years preceding diagnosis and could not assess earlier structural changes. Finally, BrainAGE quantifies deviation from normative structural ageing but does not distinguish between different biological pathways or between accelerated ageing and early neurodegenerative processes. Combining BrainAGE with molecular and fluid biomarkers will be important for clarifying these mechanisms.

A further methodological consideration is that BrainAGE was estimated independently at each imaging time point using a cross-sectional brain-age model. Although this approach allows longitudinal changes in BrainAGE to be evaluated, the model itself does not explicitly incorporate longitudinal image processing or model training. To our knowledge, fully integrated longitudinal BrainAGE models that combine longitudinal feature extraction (preprocessing) with model development remain limited because of the methodological complexity and the limited availability of sufficiently dense longitudinal imaging data. We therefore applied a validated cross-sectional BrainAGE model independently at each imaging time point, as the repeated MRI examinations required to robustly model individual ageing trajectories (for example, annual follow-up imaging) are rarely available in routine clinical practice.

Overall, our findings show that structural brain ageing begins to diverge years before the onset of cognitive impairment. This early divergence can be detected using BrainAGE and provides complementary information beyond conventional structural MRI measures. Although the improvement in prediction was modest, it was consistent across cognitive outcomes, suggesting that BrainAGE captures distributed patterns of structural vulnerability that are not fully represented by standard MRI features alone. Together, these findings support BrainAGE as a useful component of multimodal approaches for identifying cognitively normal individuals at increased risk of future cognitive decline.

## Acknowledgments

Data collection and sharing for this project was funded by the Alzheimer’s Disease Neuroimaging Initiative (ADNI) (National Institutes of Health Grant U01 AG024904) and DOD ADNI (Department of Defense award number W81XWH-12-2-0012). ADNI is funded by the National Institute on Ageing, the National Institute of Biomedical Imaging and Bioengineering, and through generous contributions from the following: AbbVie, Alzheimer’s Association; Alzheimer’s Drug Discovery Foundation; Araclon Biotech; BioClinica, Inc.; Biogen; Bristol-Myers Squibb Company; CereSpir, Inc.; Cogstate; Eisai Inc.; Elan Pharmaceuticals, Inc.; Eli Lilly and Company; EuroImmun; F. Hoffmann-La Roche Ltd and its affiliated company Genentech, Inc.; Fujirebio; GE Healthcare; IXICO Ltd.; Janssen Alzheimer Immunotherapy Research & Development, LLC.; Johnson & Johnson Pharmaceutical Research & Development LLC.; Lumosity; Lundbeck; Merck & Co., Inc.; Meso Scale Diagnostics, LLC.; NeuroRx Research; Neurotrack Technologies; Novartis Pharmaceuticals Corporation; Pfizer Inc.; Piramal Imaging; Servier; Takeda Pharmaceutical Company; and Transition Therapeutics. The Canadian Institutes of Health Research is providing funds to support ADNI clinical sites in Canada. Private sector contributions are facilitated by the Foundation for the National Institutes of Health ( https://www.fnih.org). The grantee organization is the Northern California Institute for Research and Education, and the study is coordinated by the Alzheimer’s Therapeutic Research Institute at the University of Southern California. ADNI data are disseminated by the Laboratory for Neuro Imaging at the University of Southern California.

The computational analysis was run on the servers provided by Bioinformatics Center (https://bioinformatics.uef.fi/biowhat/BIC/bic_101/), University of Eastern Finland, Finland.

During the preparation of this work, the authors used GPT-4 from OpenAI to improve language and readability. After using this tool/service, the authors reviewed and edited the content as needed. The authors take full responsibility for the content of the publication.

## Availability of data and materials

Data used in the preparation of this article were obtained from the Alzheimer’s Disease Neuroimaging Initiative (ADNI) database (https://adni.loni.usc.edu/). Details about data access are detailed there. The participant’s RIDs are provided in Supplementary Materials. Access to the OSTPRE data is restricted due to privacy considerations and required permissions.

The **Computational Anatomy Toolbox (CAT12)** used for MRI preprocessing is available at: https://github.com/ChristianGaser/cat12. The code used to generate **BrainAGE** is available at: https://github.com/ChristianGaser/BrainAGE.

The R code used for all analyses presented in this study is available at: https://github.com/ElahehMoradi/BrainAGE-EarlyCognitiveDecline

