## Supplementary Materials for "Elevated BrainAGE precedes cognitive impairment and improves prediction of future cognitive decline"

July 15, 2026

#### 1 Supplementary Methods and Materials

##### 1.1 ADNI data

In ADNI, we addressed two complementary cognitive outcomes: (i) progression from cognitively normal status to mild cognitive impairment or dementia, and (ii) longitudinal memory change, examined exploratorily through stratification into high- and low-rate decliners and predictively as a continuous outcome representing the annual rate of memory change.

For analyses of progression to MCI or dementia, we included participants who were cognitively normal or reported subjective memory complaints at baseline and who had APOE4 status, baseline composite cognitive scores, T1-weighted MRI, and longitudinal diagnostic follow-up. Participants were classified as either stable or converters based on their longitudinal diagnostic trajectories. Converters were individuals who were cognitively normal (CN) at baseline and subsequently progressed to MCI or dementia during follow-up. To reduce the likelihood of transient diagnostic fluctuations, diagnoses were required to be confirmed at the final two available visits. Stable individuals remained cognitively normal throughout follow-up and had at least five years of diagnostic follow-up to reduce the likelihood that participants classified as stable would later convert after the observation window. After applying these criteria, the final sample included 330 stable individuals and 104 converters.

For the analyses of annual memory change, we included participants with available APOE4 status, baseline composite cognitive scores, and T1 weighted MRI, who also had longitudinal composite memory scores. To enable estimation of meaningful longitudinal change, participants were required to have at least one follow-up visit occurring two years or more after baseline with an accompanying composite memory score. Participants without such follow-up data were excluded. Missing follow-up cognitive measures in ADNI primarily reflect limited follow-up duration, participant dropout, or incomplete cognitive assessments at later visits, partly due to the multi-phase structure of ADNI with variable visit schedules, in which participants do not necessarily continue across all phases or contribute late follow-up data [1]. Annual memory change was calculated as the difference between the final available follow-up and baseline composite cognitive score, divided by the time interval in years between the two assessments. For the longitudinal and cross-sectional group comparisons, participants were classified as high-rate or low-rate memory decliners using a median split of the annual rate of memory change. Individuals with an annual rate of memory decline greater than the cohort median were classified as high-rate decliners, whereas those below the median were classified as low-rate decliners.To assess potential age-related effects and ensure that this approach was not driven by baseline age, we examined the association between baseline age and memory change rate (Supplementary Fig. S1).

Longitudinal BrainAGE trajectory analyses included participants with at least two MRI scans within the predefined examination windows were included, and all available scans within these windows were used to estimate individual trajectories. Cross-sectional analyses were limited to predefined annual time points, with each participant contributing at most one MRI scan per time point.

For progression to MCI/dementia analyses, the examination window covered the four years preceding conversion to MCI or dementia. For converters, time was referenced to the visit at which conversion was first diagnosed (time $t = 0$), and MRI scans acquired during the preceding four years ($-4$ to 0 years) were included. For stable individuals, the reference point was defined as the visit occurring four years after baseline, and scans obtained during the corresponding preceding four-year period were used to maintain a comparable observation window. For memory-decline analyses, the examination window covered the four years following baseline, and MRI scans obtained within this period were used to estimate BrainAGE trajectories relative to baseline. The participant identifier list used for these analyses is available in the supplementary materials.

Baseline demographic variables, including age and sex, as well as APOE4 status and diagnostic information for the ADNI cohort, were obtained from the ADNIMERGE and DXSUM tables. Composite cognitive scores were obtained from the Phenotype Harmonization Consortium (PHC) dataset [2], provided in the file ADSP_PHC_COGN.csv. We used the memory (MEM), executive functioning (EXF), and language (LAN) composite scores. All data were downloaded from the ADNI data repository (http://adni.loni.usc.edu/).

#### 1.2 OSTPRE population-based cohort

The Kuopio Osteoporosis Risk Factor and Prevention Study (OSTPRE) is a population-based cohort that includes all women born between 1932 and 1941 who were living in Kuopio County, Eastern Finland, in 1989 (N = 14,220) [3, 4, 5, 6, 7]. Participants have been followed for three decades through repeated health surveys and linkage to national and local health registers, providing extensive longitudinal health information.

Brain MRI data for OSTPRE women were obtained from the regional picture archiving and communication system (PACS) of the public health care provider serving the region. The PACS has stored MRI examinations since 2003, routine clinical scans acquired using different scanners, field strengths, and acquisition protocols. Between 2003 and 2022, a total of 2434 T1-weighted brain MRI scans from 1885 OSTPRE participants passed quality control procedures and were available for analysis, as described previously [8]. Cognitive status at each MRI visit was determined through linkage with nationwide health registers, including hospital discharge records and other national health registers. Based on diagnostic codes recorded before the MRI visit date, participants were categorized as having no memory complaints (NMC), subjective memory complaints (SMC), mild cognitive impairment (MCI), or dementia. The OSTPRE cohort has been described in detail in earlier work [3, 4, 5, 6, 7].

For the present study, OSTPRE was used as an external replication cohort for progression to MCI/dementia analyses only. Memory decline analyses could not be replicated in OSTPRE due to the lack of harmonized longitudinal cognitive composite measures. For progression analyses, we included participants who were cognitively normal at their first available MRI visit, defined as having no memory complaints or only subjective memory complaints at that visit. This first MRI served as the baseline examination, and follow-up diagnoses were determined from register data recorded after baseline. Because MRI examinations in OSTPRE were obtained from routine clinical practice rather than specialized memory clinic settings, participants underwent brain imaging for a wide range of non–cognitive indications (e.g., headache, dizziness, trauma evaluation, or vascular assessment). Thus, undergoing MRI at baseline does not imply suspected cognitive decline, and the clinical indications leading to imaging are heterogeneous and not systematically related to neurodegenerative disease. Register-based cognitive status at the MRI visit further ensured that all included participants were cognitively normal at baseline (at the first MRI visit).

Participants were classified as converters if they subsequently received a register-based diagnosis of MCI or dementia during follow-up, and as stable if no such diagnosis was recorded during the available observation period. To account for potential delays between underlying disease onset and the recorded diagnosis date, diagnostic states were assumed to extend up to one year prior to the registered diagnosis. Accordingly, the estimated conversion time reflects the earliest time point within this window rather than the recorded diagnosis date. Because OSTPRE is a population-based cohort with variable follow-up durations, participants without a recorded diagnosis were classified as stable based on the available observation period. However, some individuals may have had insufficient follow-up time for a diagnosis to emerge or may have died before a diagnosis was recorded. To reduce potential misclassification, we restricted the stable group to participants with at least five years of follow-up after the baseline MRI or until the end of the study period (2022). Participants who died or reached the end of follow-up before completing five years without a recorded diagnosis were excluded from the stable group.

For longitudinal BrainAGE trajectory analyses, we included participants with at least two MRI scans acquired within the predefined examination window. All available scans from these individuals within this period were used to estimate individual BrainAGE trajectories. To assess potential selection bias associated with requiring repeated MRI scans, we compared participants with and without multiple scans within this window in terms of conversion status and BrainAGE measures (Supplementary Table S1). Participants with repeated scans showed a slightly higher proportion of converters, while BrainAGE distributions were comparable across groups. These findings suggest that any resulting selection bias is limited and unlikely to materially influence the results. For cross-sectional analyses (aligned to the time of conversion), MRI scans were not obtained at fixed time points in OSTPRE, unlike the more regular follow-up schedule in ADNI. Therefore, scans were assigned to the nearest annual time point. When multiple scans fell within the same one-year interval, the scan closest to the target time point was selected.

The longitudinal data structure differed between ADNI and OSTPRE. In ADNI, participants were followed within a structured research protocol with regular visits, resulting in a higher number of longitudinal MRI observations per individual. In contrast, in OSTPRE, fewer participants contributed repeated MRI data (85 stable-CN and 56 converter-CN vs. 281 stable-CN and 81 converter-CN in ADNI), and MRI acquisitions were less frequent and not systematically timed. In addition, models in ADNI were adjusted for sex and APOE4 status, whereas such adjustments were not possible in OSTPRE, as all participants were women and APOE4 data were not available. These differences are important to consider when interpreting cross-cohort comparisons.

#### 1.3 Additional details of statistical analyses

A four-year time window was used for all longitudinal analyses. For progression analyses, time was defined relative to the conversion event, with time 0 corresponding to the first diagnosis of MCI or dementia in converters and negative values representing years prior to conversion ($-4$ to 0 years). For stable CN participants who did not convert during follow-up, an analogous time axis was defined relative to baseline, with the baseline visit assigned to the $-4$ year timepoint and subsequent follow-up visits used to approximate the interval from $-4$ to 0 years, ensuring comparable observation windows between groups. For memory-decline analyses, time was defined relative to baseline, covering the interval from baseline to four years after baseline (0 to 4 years).

Participants were included in longitudinal analyses if they had at least two MRI scans within the relevant time window. All available scans within these windows were used to estimate individual BrainAGE trajectories. In ADNI, models were adjusted for APOE4 status and sex. In OSTPRE, APOE4 was not available, and sex was not included because all participants were women. A random intercept term was included to account for repeated measurements. Models were estimated using restricted maximum likelihood (REML), and two-sided $P$-values were obtained using Wald t-tests with Satterthwaite approximations for the degrees of freedom [9]. These analyses were performed using the `lmerTest` package in R [10].

For cross-sectional analyses of progression to MCI or dementia, time points were defined at 4, 3, 2, 1, and 0 years before conversion, with time 0 corresponding to the first diagnosis of MCI or dementia in converters. For stable cognitively normal participants, timepoints were defined relative to baseline, with the baseline visit corresponding to the $-4$ year timepoint and subsequent follow-up visits approximating the $-3$, $-2$, $-1$, and 0 year timepoints. For memory-decline analyses, timepoints were defined at 0, 1, 2, 3, and 4 years after baseline.

At each timepoint, BrainAGE values were derived from the MRI scan closest to the target year. Because MRI acquisition schedules differed across individuals, sample sizes varied across timepoints and BrainAGE values varied within individuals across time. Each model included only one observation per participant at a given timepoint. In ADNI, cross-sectional models were adjusted for APOE $\varepsilon4$ status and sex. Discriminative performance was evaluated using receiver operating characteristic (ROC) analysis, with BrainAGE as the predictor and group status as the outcome, and summarized using the area under the curve (AUC).

As a sensitivity analysis, all longitudinal and cross-sectional models were repeated in a restricted subset of participants who had MRI-derived BrainAGE measures available at predefined timepoints ($-4$, $-2$, and 0 years). Only these timepoints were included in the analysis, and all other observations were excluded. This restriction ensured consistent temporal alignment across individuals and allowed us to assess whether the results were robust to variability in follow-up timing and scan availability.

For the time-to-event analyses, BrainAGE was dichotomized into BrainAGE-positive (BA+) and BrainAGE-negative (BA-) groups, defined as values greater than or less than zero, respectively. Time was measured from the baseline visit to the first occurrence of MCI or dementia, with participants who did not convert during follow-up censored at their last available diagnostic assessment and deaths treated as censoring events. The proportional hazards assumption was evaluated using Schoenfeld residuals [11], with $P < 0.05$ indicating violation. Cumulative incidence of conversion was additionally estimated while accounting for death as a competing risk using the Aalen–Johansen estimator, and differences between groups were assessed using Gray's test [12]. Analyses were performed separately for Global and Temporal BrainAGE using the `survival` [13] and `cmprsk` [14] packages in R.

#### 1.4 Additional details of predictive modelling

The predictive framework followed our previously described approach [15]. Because structural MRI measures (regional volumes and cortical thickness) were substantially higher-dimensional than the remaining predictors, a two-stage framework was used to prevent imaging features from dominating the predictive models. In the first stage, baseline MRI measures were reduced to a single task-specific MRI-derived score using elastic net regularisation [16]. Separate MRI-derived scores were generated for each outcome (conversion to MCI or dementia and annual change in memory performance), capturing multivariate MRI patterns associated with the outcome of interest. In the second stage, these MRI-derived scores were combined with BrainAGE measures and non-imaging predictors.

Task-specific elastic net models were then fitted using logistic regression for conversion analyses and linear regression for prediction of annual change in memory performance. Model performance was evaluated using nested, stratified 10-fold cross-validation repeated 10 times. The inner cross-validation loop was used to derive the MRI-derived score, whereas the outer loop provided independent training and test sets for model evaluation, ensuring complete separation between feature derivation and performance assessment.

Classification performance was evaluated using the area under the receiver operating characteristic curve (AUC). Prediction of annual memory change was evaluated using Pearson correlation and mean absolute error (MAE). Performance measures were averaged across repeated cross-validation runs. Additional classification metrics, including balanced accuracy, sensitivity, and specificity, are reported in the Supplementary Results.

Pairwise comparisons of AUC values were performed using DeLong's test on the median-performing cross-validation run. Correlation coefficients were compared using the Hittner modification of the Dunn–Clark test [17, 18] on the cross-validation run with the median correlation, implemented using the `CoCor` package [19].

These analyses were performed using the `glmnet` [20], `caret` [21], `pROC` [22], and `CoCor` [19] packages in R (version 4.1.1). Data preparation was performed using `dplyr` [23], and visualisations were generated using `ggplot2` [24] and `ComplexHeatmap` [25].

[20] Trevor Hastie, Junyang Qian, and Kenneth Tay. An introduction to glmnet. *CRAN R Repositary*, 2021.

### 2 Supplementary Tables

Table S1: Comparison of participants with ≥2 MRI scans within the 4-year window preceding conversion (or equivalent reference time for stable individuals) (Group 1) and those with less than 2 MRIs in this time window (Group 2) in OSTPRE. Values are proportions or mean (standard deviation) of BrainAGE measures within converters (CCN) and stable individuals (SCN).

| | **Group 1: ≥2 MRIs** | **Group 2: <2 MRIs** |
|---|---|---|
| *Sample size* | | |
| Total $n$ | 141 | 646 |
| Converters (CCN), $n$ (%) | 56 (39.7%) | 210 (32.5%) |
| Stable (SCN), $n$ (%) | 85 (60.3%) | 436 (67.5%) |
| *Global BrainAGE, mean (standard deviation)* | | |
| Converters (CCN) | 0.25 (4.34) | 0.78 (3.75) |
| Stable (SCN) | -1.99 (4.88) | -2.26 (4.11) |
| *Temporal BrainAGE, mean(standard deviation)* | | |
| Converters (CCN) | 0.10 (4.96) | 1.00 (4.62) |
| Stable (SCN) | -2.96 (4.97) | -2.80 (4.87) |

Table S2: Longitudinal mixed-effects models of BrainAGE in ADNI (subjects with ≥2 visits).

| Effect | Global BrainAGE | | Temporal BrainAGE | |
|---|---|---|---|---|
| | Estimate (95% CI) | $t$ / $p$ | Estimate (95% CI) | $t$ / $p$ |
| Time (years) | 0.11 [0.07, 0.15] | $t = 5.55$, $p = 3.7 \times 10^{-8}$ | 0.21 [0.17, 0.24] | $t = 10.80$, $p < 2 \times 10^{-16}$ |
| Group (Converter vs Stable) | 3.29 [2.43, 4.15] | $t = 7.45$, $p = 5.8 \times 10^{-13}$ | 3.37 [2.48, 4.26] | $t = 7.39$, $p = 9.0 \times 10^{-13}$ |
| Time × Group | 0.17 [0.07, 0.26] | $t = 3.49$, $p = 5.1 \times 10^{-4}$ | 0.23 [0.14, 0.33] | $t = 5.01$, $p = 6.2 \times 10^{-7}$ |
| APOE4 | -0.17 [-0.88, 0.55] | $t = -0.45$, $p = 0.650$ | -0.31 [-1.05, 0.43] | $t = -0.82$, $p = 0.412$ |
| Sex (Male) | 0.35 [-0.36, 1.05] | $t = 0.97$, $p = 0.335$ | 0.59 [-0.14, 1.32] | $t = 1.57$, $p = 0.117$ |

Table S3: Cross-sectional group differences and discriminative performance of Global and Temporal BrainAGE at timepoints preceding diagnosis of MCI/dementia in ADNI. $\beta$ denotes the adjusted regression coefficient (Converter–Stable) from linear models controlling for APOE4 status and sex. AUC values reflect univariate discrimination using BrainAGE alone.

| Outcome | Years | $\beta$ | 95% CI | $p$ value | AUC | 95% CI | $n_{\text{Stable}}/n_{\text{Converter}}$ |
|---|---|---|---|---|---|---|---|
| **Global BrainAGE** | −4 | 2.83 | [1.80, 3.87] | $1.3 \times 10^{-7}$ | 0.72 | [0.63, 0.81] | 330 / 45 |
| | −3 | 3.30 | [2.14, 4.46] | $6.5 \times 10^{-8}$ | 0.75 | [0.67, 0.82] | 183 / 40 |
| | −2 | 3.43 | [2.37, 4.48] | $7.6 \times 10^{-10}$ | 0.73 | [0.66, 0.81] | 226 / 53 |
| | −1 | 2.65 | [1.26, 4.04] | $2.7 \times 10^{-4}$ | 0.69 | [0.59, 0.79] | 66 / 44 |
| | 0 | 3.19 | [1.95, 4.42] | $8.7 \times 10^{-7}$ | 0.72 | [0.64, 0.80] | 127 / 53 |
| **Temporal BrainAGE** | −4 | 3.09 | [2.05, 4.13] | $1.0 \times 10^{-8}$ | 0.73 | [0.66, 0.81] | 330 / 45 |
| | −3 | 3.06 | [1.89, 4.24] | $6.4 \times 10^{-7}$ | 0.73 | [0.65, 0.81] | 183 / 40 |
| | −2 | 3.41 | [2.31, 4.50] | $3.2 \times 10^{-9}$ | 0.72 | [0.64, 0.80] | 226 / 53 |
| | −1 | 2.24 | [0.80, 3.68] | $2.6 \times 10^{-3}$ | 0.65 | [0.55, 0.75] | 66 / 44 |
| | 0 | 2.90 | [1.63, 4.16] | $1.1 \times 10^{-5}$ | 0.68 | [0.60, 0.77] | 127 / 53 |

Table S4: Sensitivity analysis: longitudinal mixed-effects models of BrainAGE in a restricted ADNI subset with MRI data available at all selected timepoints preceding diagnosis of MCI/dementia (−4, −2, and 0 years; 15 converters, 117 stables).

| Effect | Global BrainAGE | | Temporal BrainAGE | |
|---|---|---|---|---|
| | Estimate (95% CI) | $t$ / $p$ | Estimate (95% CI) | $t$ / $p$ |
| Time (years) | 0.10 [0.04, 0.16] | $t = 3.49$, $p = 0.0006$ | 0.20 [0.15, 0.26] | $t = 7.57$, $p = 6.22 \times 10^{-13}$ |
| Group (Converter vs Stable) | 4.30 [2.41, 6.19] | $t = 4.50$, $p = 1.41 \times 10^{-5}$ | 3.70 [1.81, 5.59] | $t = 3.81$, $p = 0.0002$ |
| Time × Group | 0.30 [0.13, 0.47] | $t = 3.47$, $p = 0.0006$ | 0.32 [0.16, 0.48] | $t = 4.03$, $p = 7.40 \times 10^{-5}$ |
| APOE4 | −0.06 [−1.31, 1.20] | $t = -0.09$, $p = 0.930$ | −0.68 [−1.94, 0.58] | $t = -1.06$, $p = 0.294$ |
| Sex (Male) | 0.26 [−0.90, 1.42] | $t = 0.44$, $p = 0.660$ | 0.59 [−0.58, 1.76] | $t = 0.98$, $p = 0.330$ |

Table S5: Sensitivity analysis: cross-sectional group differences and discriminative performance of BrainAGE in a restricted ADNI subset with MRI data available at all selected timepoints preceding diagnosis of MCI/dementia (−4, −2, and 0 years; 15 converters, 117 stables). $\beta$ denotes the adjusted regression coefficient (Converter vs Stable) from linear models controlling for APOE4status and sex. AUC values reflect univariate discrimination using BrainAGE alone.

| Years to conversion | Global BrainAGE | | Temporal BrainAGE | |
|---|---|---|---|---|
| | $\beta$ (95% CI) | AUC (95% CI) | $\beta$ (95% CI) | AUC (95% CI) |
| −4 | 3.10 [1.24, 4.96] | 0.74 [0.61, 0.86] | 2.40 [0.51, 4.29] | 0.68 [0.55, 0.81] |
| −2 | 3.65 [1.70, 5.60] | 0.75 [0.63, 0.88] | 3.03 [1.09, 4.97] | 0.72 [0.59, 0.84] |
| 0 | 4.34 [2.36, 6.32] | 0.78 [0.66, 0.90] | 3.74 [1.77, 5.71] | 0.74 [0.62, 0.86] |

Table S6: Longitudinal mixed-effects models of Global and Temporal BrainAGE in the OSTPRE cohort. Models included time to conversion, diagnostic group (Converter vs Stable), and their interaction, with a random intercept for subject.

| Effect | Global BrainAGE | | Temporal BrainAGE | |
|---|---|---|---|---|
| | Estimate (95% CI) | $t$ / $p$ | Estimate (95% CI) | $t$ / $p$ |
| Time (years) | 0.17 [−0.11, 0.45] | $t = 1.18$, $p = 0.241$ | 0.33 [0.05, 0.62] | $t = 2.27$, $p = 0.024$ |
| Group (Converter vs Stable) | 3.76 [1.84, 5.69] | $t = 3.83$, $p = 0.0002$ | 3.96 [1.99, 5.93] | $t = 3.92$, $p = 0.0001$ |
| Time × Group | 0.37 [−0.12, 0.86] | $t = 1.50$, $p = 0.134$ | 0.16 [−0.34, 0.66] | $t = 0.62$, $p = 0.537$ |

Table S7: Cross-sectional group differences and discriminative performance of Global and Temporal BrainAGE at timepoints preceding diagnosis of MCI/dementia in the OSTPRE cohort. $\beta$ denotes the regression coefficient (Converter–Stable) from cross-sectional linear models. AUC values reflect univariate discrimination using BrainAGE alone.

| Outcome | Years | $\beta$ | 95% CI | $p$ | AUC | 95% CI | $n_{\text{Stable}}/n_{\text{Converter}}$ |
|---|---|---|---|---|---|---|---|
| **Global BrainAGE** | $-4$ | 2.76 | [1.51, 4.01] | $< 10^{-4}$ | 0.69 | [0.61, 0.77] | 521 / 48 |
| | $-3$ | 3.53 | [1.74, 5.33] | $< 10^{-3}$ | 0.71 | [0.60, 0.82] | 35 / 71 |
| | $-2$ | 2.11 | [0.44, 3.79] | 0.014 | 0.66 | [0.53, 0.78] | 33 / 83 |
| | $-1$ | 3.26 | [1.33, 5.19] | 0.001 | 0.70 | [0.57, 0.82] | 27 / 97 |
| | 0 | 2.92 | [1.04, 4.80] | 0.003 | 0.68 | [0.55, 0.80] | 26 / 65 |
| **Temporal BrainAGE** | $-4$ | 3.40 | [1.96, 4.83] | $< 10^{-4}$ | 0.71 | [0.63, 0.78] | 521 / 48 |
| | $-3$ | 4.58 | [2.54, 6.61] | $< 10^{-4}$ | 0.74 | [0.64, 0.83] | 35 / 71 |
| | $-2$ | 3.02 | [1.10, 4.94] | 0.002 | 0.69 | [0.58, 0.80] | 33 / 83 |
| | $-1$ | 5.81 | [3.85, 7.78] | $< 10^{-6}$ | 0.79 | [0.68, 0.89] | 27 / 97 |
| | 0 | 4.62 | [2.69, 6.55] | $< 10^{-5}$ | 0.75 | [0.64, 0.87] | 26 / 65 |

Table S8: Cause-specific Cox regression analysis of conversion from cognitively normal status to MCI/dementia. Hazard ratios (HRs) with 95% confidence intervals (CIs) were estimated for BrainAGE group (BA+/BA-). Conversion to MCI/dementia was treated as the event of interest, while death was treated as a censoring event. The proportional hazards assumption was assessed using Schoenfeld residuals.

| Cohort | BrainAGE measure | $n$ | Conversion events | HR (95% CI) | $p$ |
|---|---|---|---|---|---|
| ADNI | Global BrainAGE | 434 | 104 | 1.98 [1.34, 2.91] | $4 \times 10^{-4}$ |
| | Temporal BrainAGE | 434 | 104 | 1.60 [1.09, 2.36] | 0.020 |
| OSTPRE | Global BrainAGE | 908 | 387 | 2.58 [2.11, 3.16] | $< 2 \times 10^{-16}$ |
| | Temporal BrainAGE | 908 | 387 | 3.05 [2.49, 3.73] | $< 2 \times 10^{-16}$ |

Table S9: Longitudinal mixed-effects models of Global and Temporal BrainAGE by rate of memory decline in ADNI.

| Effect | Global BrainAGE | | Temporal BrainAGE | |
|---|---|---|---|---|
| | Estimate (95% CI) | $t$ / $p$ | Estimate (95% CI) | $t$ / $p$ |
| Time (years) | 0.12 [0.10, 0.15] | $t = 9.63$, $p < 2 \times 10^{-16}$ | 0.22 [0.20, 0.25] | $t = 17.57$, $p < 2 \times 10^{-16}$ |
| Group (High vs Low rate) | 2.04 [1.64, 2.44] | $t = 10.03$, $p < 2 \times 10^{-16}$ | 2.54 [2.12, 2.97] | $t = 11.77$, $p < 2 \times 10^{-16}$ |
| Time × Group | 0.31 [0.27, 0.35] | $t = 17.08$, $p < 2 \times 10^{-16}$ | 0.37 [0.33, 0.40] | $t = 20.60$, $p < 2 \times 10^{-16}$ |
| APOE $\varepsilon$4 | 0.39 [0.07, 0.70] | $t = 2.38$, $p = 0.017$ | 0.26 [-0.07, 0.60] | $t = 1.54$, $p = 0.124$ |
| Sex (Male) | 0.64 [0.26, 1.03] | $t = 3.27$, $p = 0.0011$ | 0.75 [0.34, 1.16] | $t = 3.59$, $p = 0.0003$ |

Table S10: Cross-sectional group differences and discriminative performance of Global and Temporal BrainAGE by rate of memory decline (high vs low) in ADNI. $\beta$ denotes the adjusted regression coefficient (high-rate vs low-rate) controlling for APOE4 status and sex.

| Outcome | Years | $\beta$ | 95% CI | $p$ value | AUC | 95% CI | $n_{\text{High decline}}/n_{\text{Low decline}}$ |
|---|---|---|---|---|---|---|---|
| **Global BrainAGE** | 0 | 2.11 | [1.73, 2.49] | $8.8 \times 10^{-27}$ | 0.67 | [0.64, 0.70] | 717 / 719 |
| | 1 | 2.47 | [2.03, 2.91] | $2.5 \times 10^{-27}$ | 0.69 | [0.66, 0.72] | 602 / 535 |
| | 2 | 2.64 | [2.18, 3.09] | $1.4 \times 10^{-28}$ | 0.70 | [0.67, 0.73] | 566 / 564 |
| | 3 | 3.15 | [2.46, 3.83] | $4.8 \times 10^{-18}$ | 0.75 | [0.70, 0.79] | 271 / 227 |
| | 4 | 3.24 | [2.52, 3.96] | $1.2 \times 10^{-17}$ | 0.73 | [0.68, 0.77] | 256 / 252 |
| **Temporal BrainAGE** | 0 | 2.56 | [2.16, 2.96] | $4.7 \times 10^{-34}$ | 0.69 | [0.66, 0.71] | 717 / 719 |
| | 1 | 3.03 | [2.56, 3.49] | $1.8 \times 10^{-35}$ | 0.72 | [0.69, 0.75] | 602 / 535 |
| | 2 | 3.41 | [2.93, 3.89] | $2.9 \times 10^{-41}$ | 0.74 | [0.71, 0.77] | 566 / 564 |
| | 3 | 3.68 | [2.95, 4.41] | $2.8 \times 10^{-21}$ | 0.76 | [0.72, 0.81] | 271 / 227 |
| | 4 | 3.97 | [3.24, 4.71] | $7.3 \times 10^{-24}$ | 0.76 | [0.72, 0.81] | 256 / 252 |

Table S11: Sensitivity analysis in ADNI: Longitudinal mixed-effects models of global and temporal BrainAGE by rate of memory decline (participants with complete follow-up at 0, 2, and 4 years). (updated)

| Effect | Global BrainAGE | | Temporal BrainAGE | |
|---|---|---|---|---|
| | Estimate (95% CI) | $p$ value | Estimate (95% CI) | $p$ value |
| Time (years) | 0.13 [0.08, 0.17] | $p = 1.08 \times 10^{-7}$ | 0.22 [0.18, 0.27] | $p < 2 \times 10^{-16}$ |
| High-rate vs Low-rate decline | 2.15 [1.47, 2.83] | $p = 1.46 \times 10^{-9}$ | 2.73 [2.03, 3.43] | $p = 1.28 \times 10^{-13}$ |
| Time × Group | 0.29 [0.22, 0.35] | $p < 2 \times 10^{-16}$ | 0.35 [0.29, 0.42] | $p < 2 \times 10^{-16}$ |
| APOE $\varepsilon 4$ | 0.46 [-0.07, 0.98] | $p = 0.088$ | 0.35 [-0.18, 0.89] | $p = 0.199$ |
| Sex (Male) | 0.56 [-0.09, 1.20] | $p = 0.092$ | 0.49 [-0.18, 1.15] | $p = 0.151$ |

Table S12: Sensitivity analysis (ADNI): Cross-sectional group differences and discriminative performance of global and temporal BrainAGE by rate of memory decline.

| Outcome | Time (years) | $\beta$ (High–Low) | 95% CI | $p$ value | AUC |
|---|---|---|---|---|---|
| Global BrainAGE | 0 | 2.27 | [1.62, 2.91] | $< 10^{-10}$ | 0.69 |
| | 2 | 2.67 | [1.98, 3.36] | $< 10^{-12}$ | 0.70 |
| | 4 | 3.24 | [2.50, 3.97] | $< 10^{-16}$ | 0.73 |
| Temporal BrainAGE | 0 | 2.83 | [2.16, 3.50] | $< 10^{-15}$ | 0.71 |
| | 2 | 3.43 | [2.73, 4.14] | $< 10^{-19}$ | 0.74 |
| | 4 | 4.06 | [3.30, 4.81] | $< 10^{-23}$ | 0.77 |

### 3 Supplementary Figures

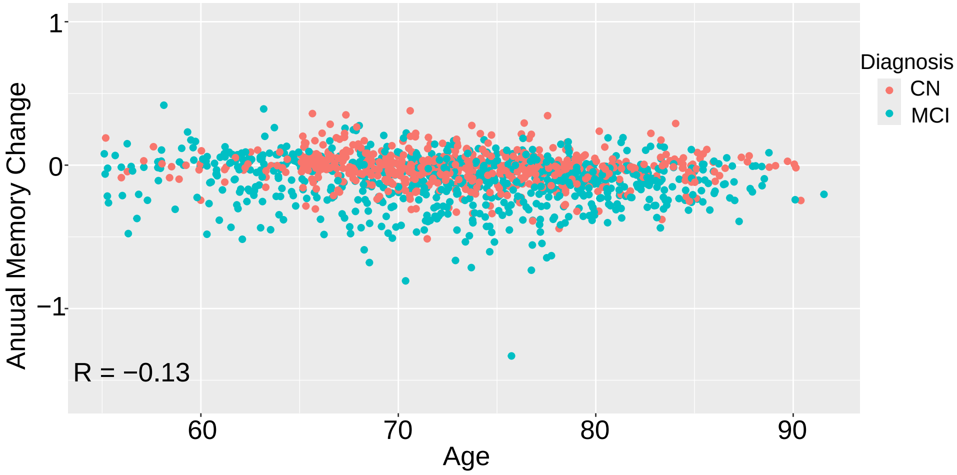


Figure S1: Relationship between baseline age and individual annual memory change, calculated as the change in composite memory score divided by the follow-up interval in ADNI. This analysis was performed to assess potential age-related effects on the annual memory change measure. No strong association was observed between baseline age and annual memory change. MCI, mild cognitive impairment; CN, cognitively normal.

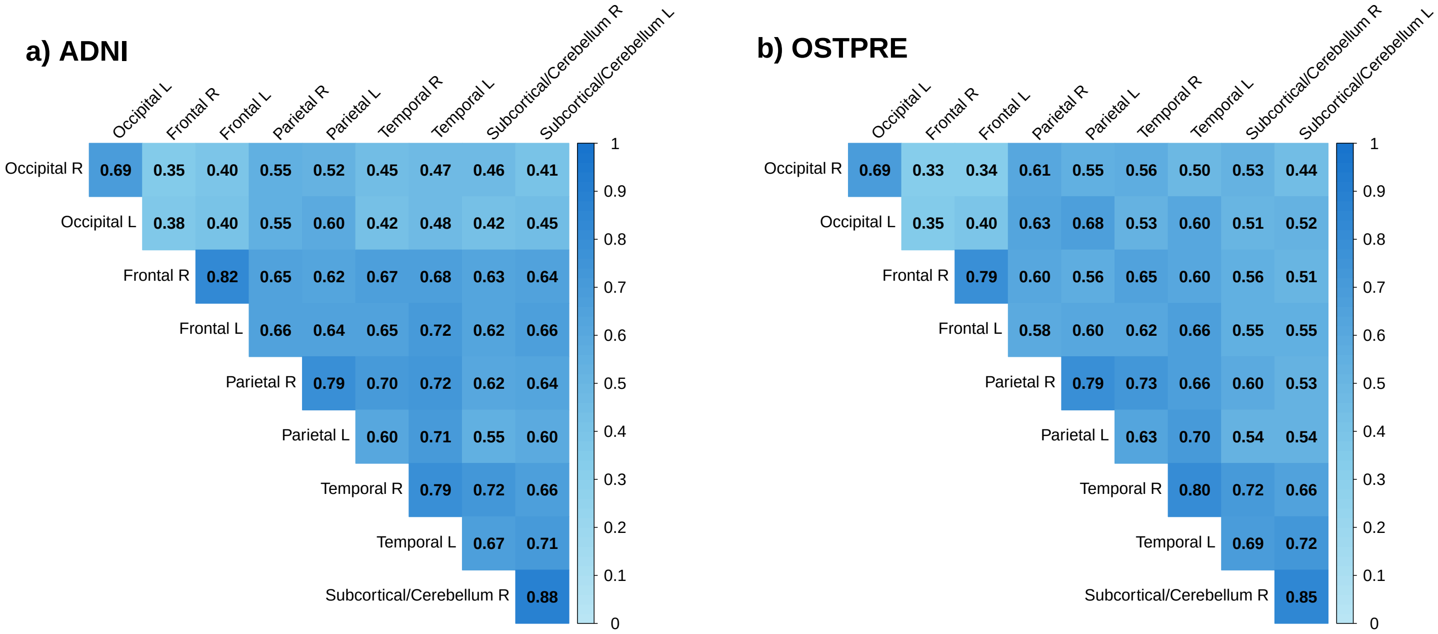


Figure S2: Correlation matrix of regional BrainAGE estimates in the ADNI and OSTPRE cohorts. Pearson correlation coefficients (range 0–1) are shown between regional BrainAGE measures in the left and right hemispheres. As expected, corresponding regions in the left and right hemispheres showed strong correlations in both cohorts, supporting the use of hemisphere-averaged regional BrainAGE measures in subsequent analyses.

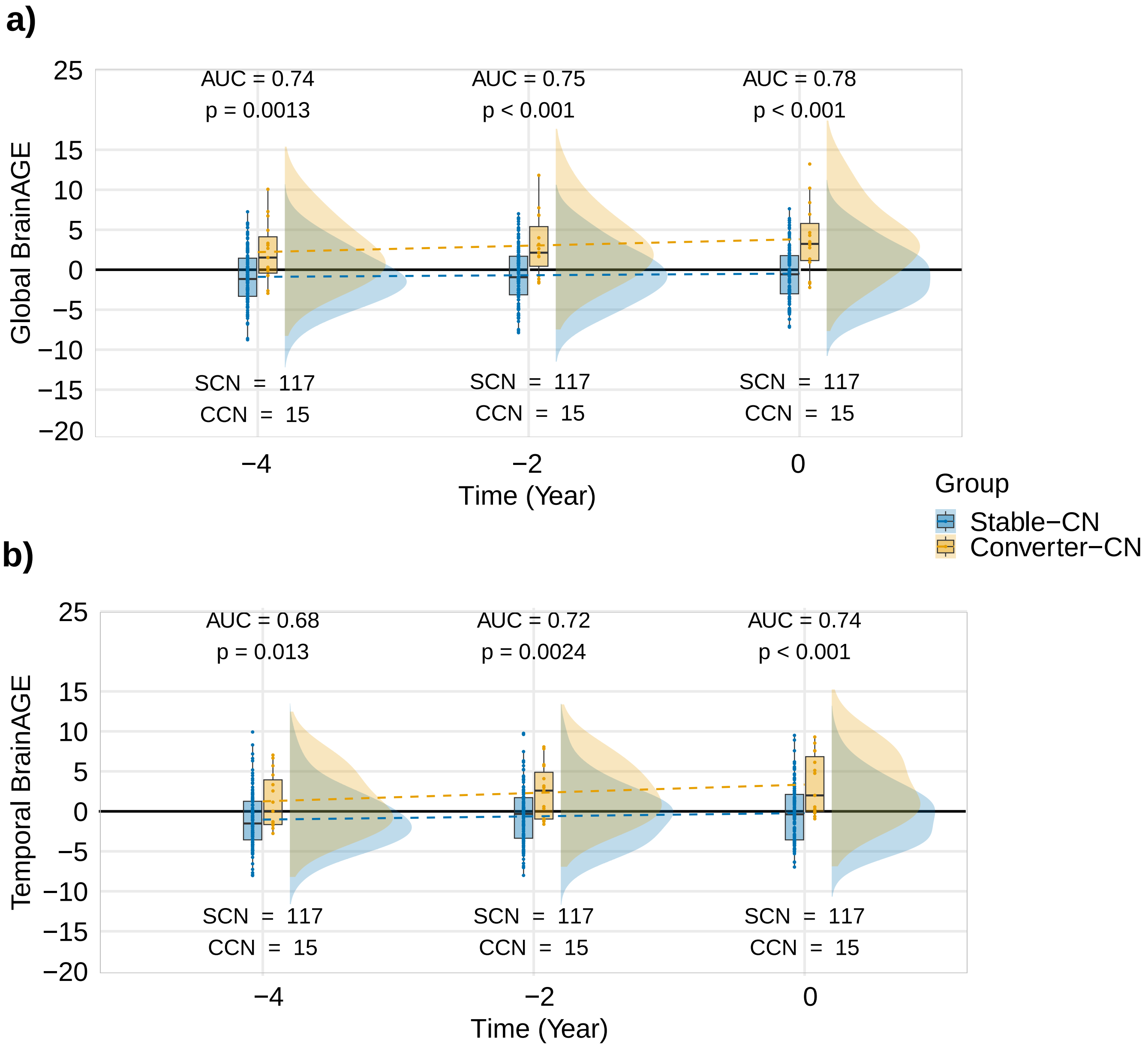


Figure S3: Longitudinal BrainAGE patterns preceding diagnosis of MCI or dementia in ADNI. Raincloud plots show BrainAGE values aligned to the time of diagnosis among cognitively normal individuals who remained stable or later converted, restricted to participants with complete longitudinal data across all examined time points. Panels display (a) Global BrainAGE and (b) Temporal BrainAGE. Dashed lines indicate group-level trends estimated using linear regression fitted separately for each group. Sample sizes and univariate AUC values are shown at each time point. These plots demonstrate consistently higher BrainAGE values and steeper longitudinal increases in individuals who subsequently converted to MCI or dementia.

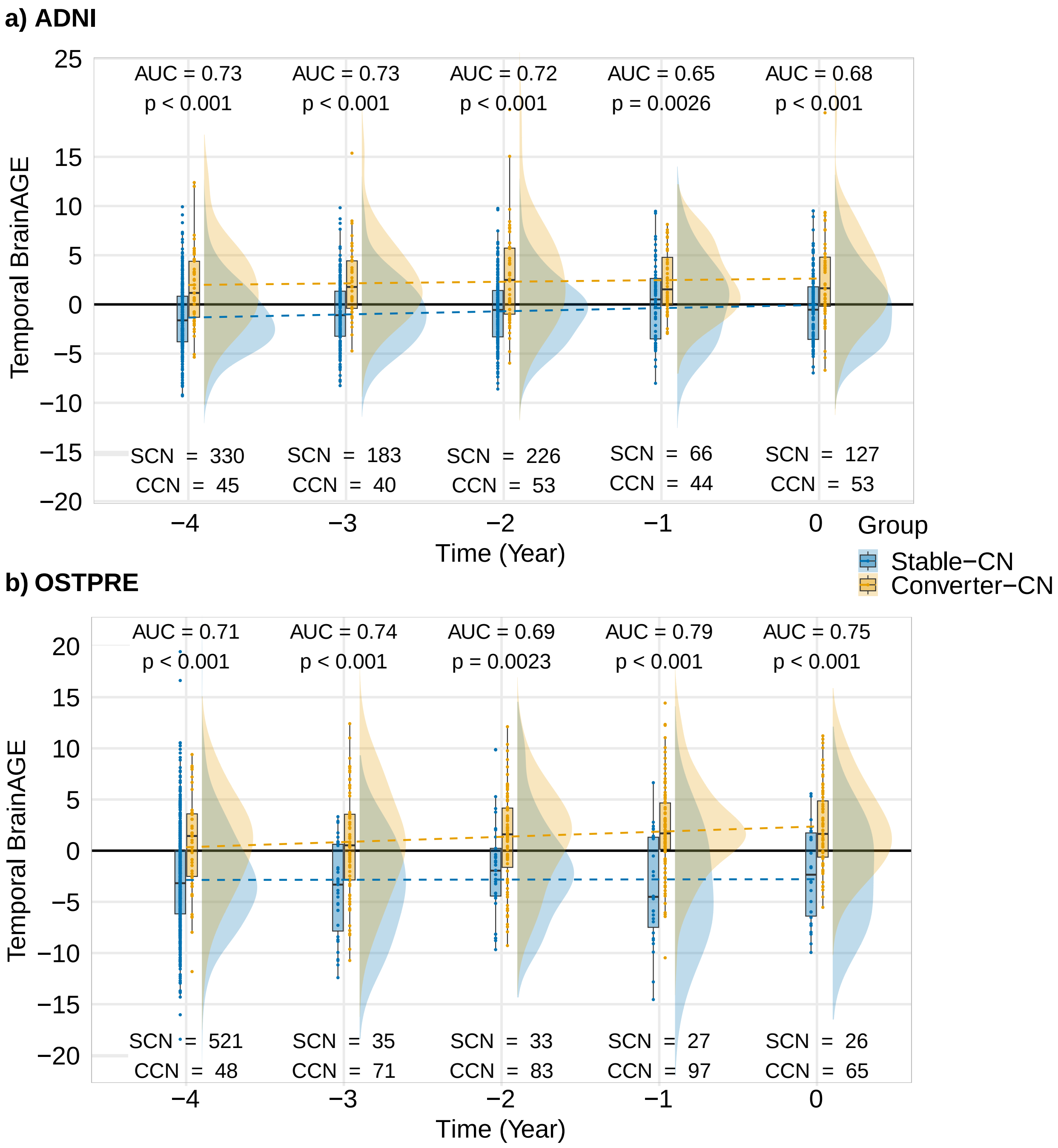


Figure S4: Longitudinal Temporal BrainAGE patterns preceding diagnosis of MCI or dementia in ADNI and OSTPRE. Raincloud plots show Temporal BrainAGE values aligned to the time of diagnosis among cognitively normal individuals who remained stable or later converted (stable-CN and converter-CN) in ADNI (a) and OSTPRE (b). Dashed lines indicate group-level trends estimated using linear regression fitted separately for each group. Sample sizes and univariate AUC values are shown at each time point. These plots show consistently higher Temporal BrainAGE values in converters across the examined time points in both cohorts.

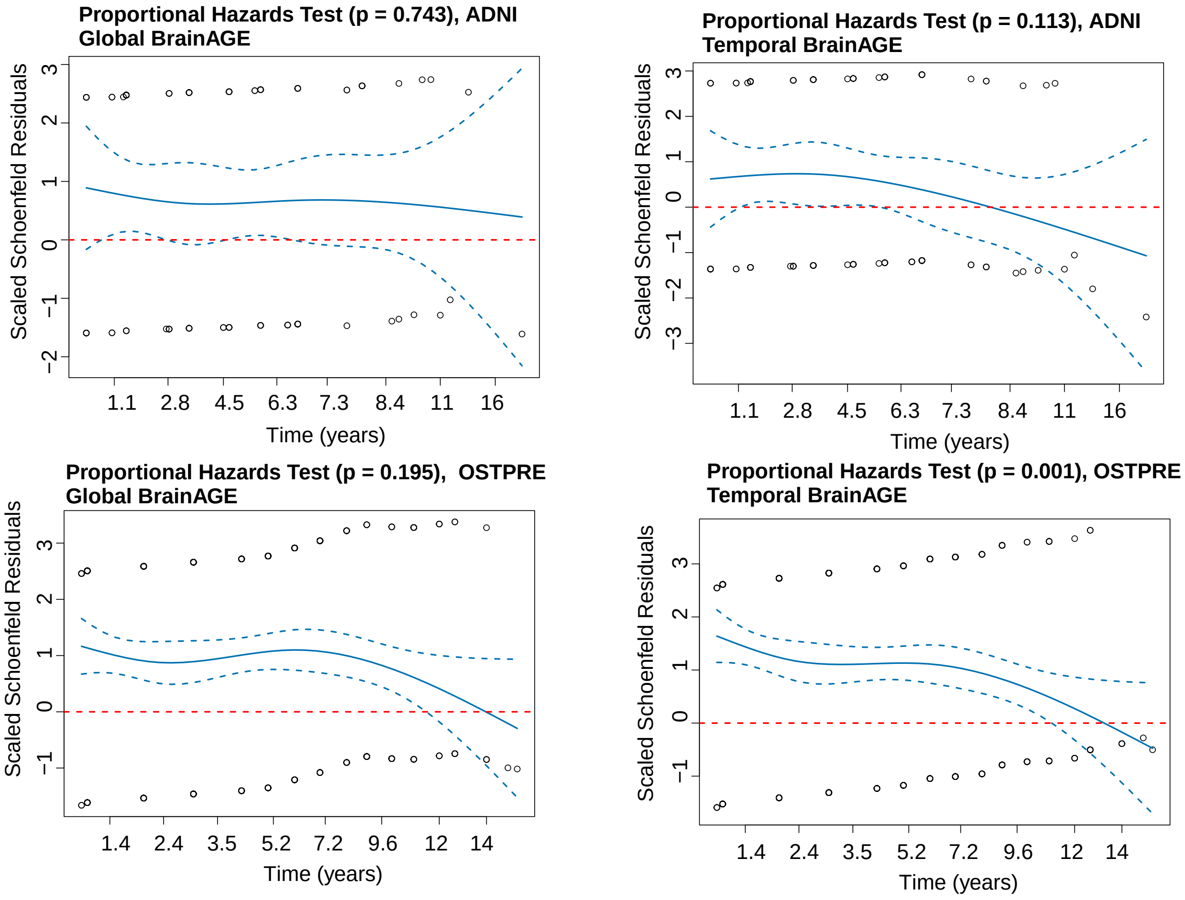


Figure S5: Evaluation of the proportional hazards assumption using Schoenfeld residuals for Cox regression models in ADNI and OSTPRE. Scaled Schoenfeld residuals are plotted against time for Global and Temporal BrainAGE predictors. The proportional hazards assumption was satisfied for all models except Temporal BrainAGE in OSTPRE, which showed evidence of time-dependent effects.

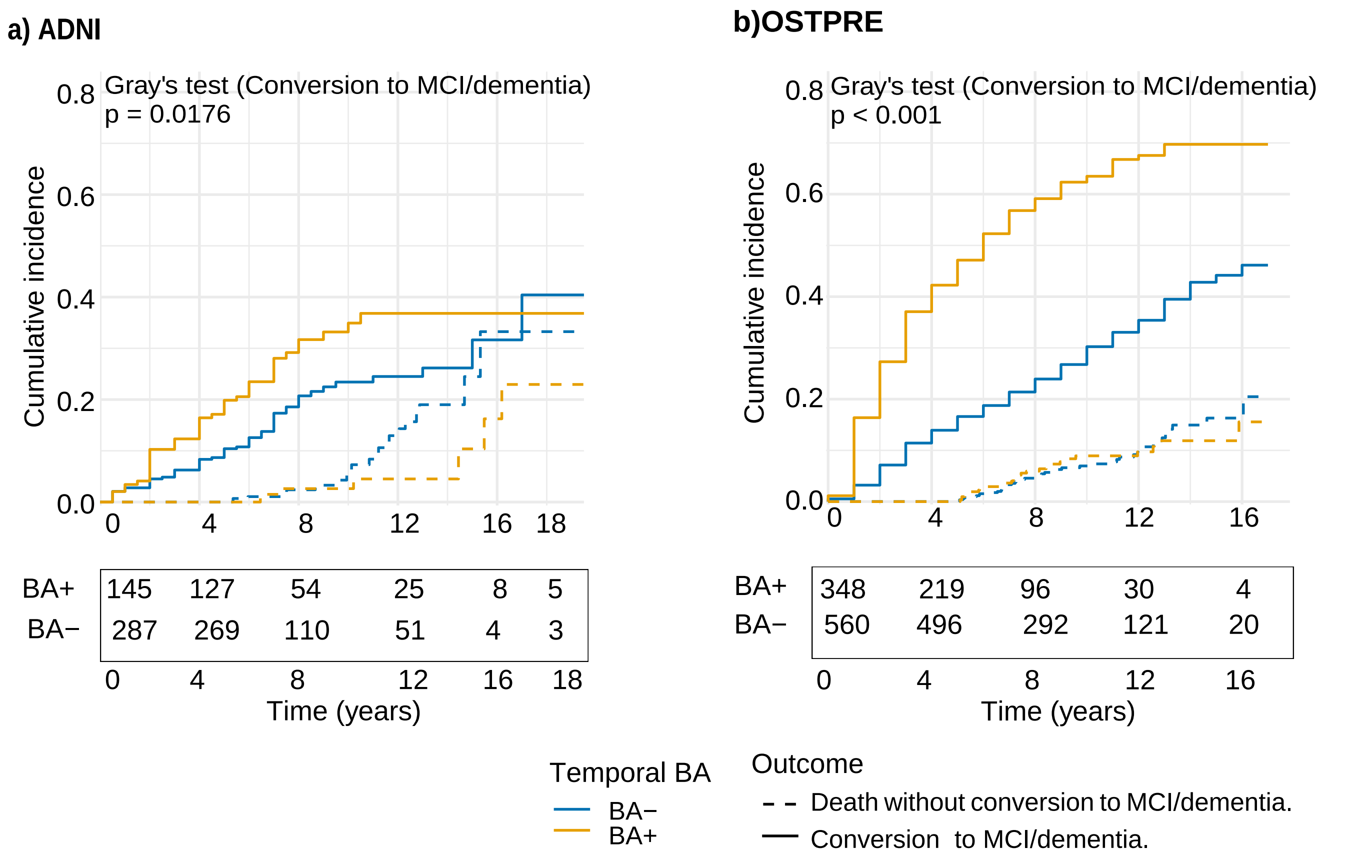


Figure S6: Cumulative incidence of conversion to MCI or dementia among cognitively normal (CN) individuals, accounting for death as a competing risk, stratified by Temporal BrainAGE (BA+/BA) in ADNI and OSTPRE. Individuals with positive Temporal BrainAGE (BA+) showed a higher cumulative incidence of conversion than those with negative Temporal BrainAGE (BA) in both cohorts.

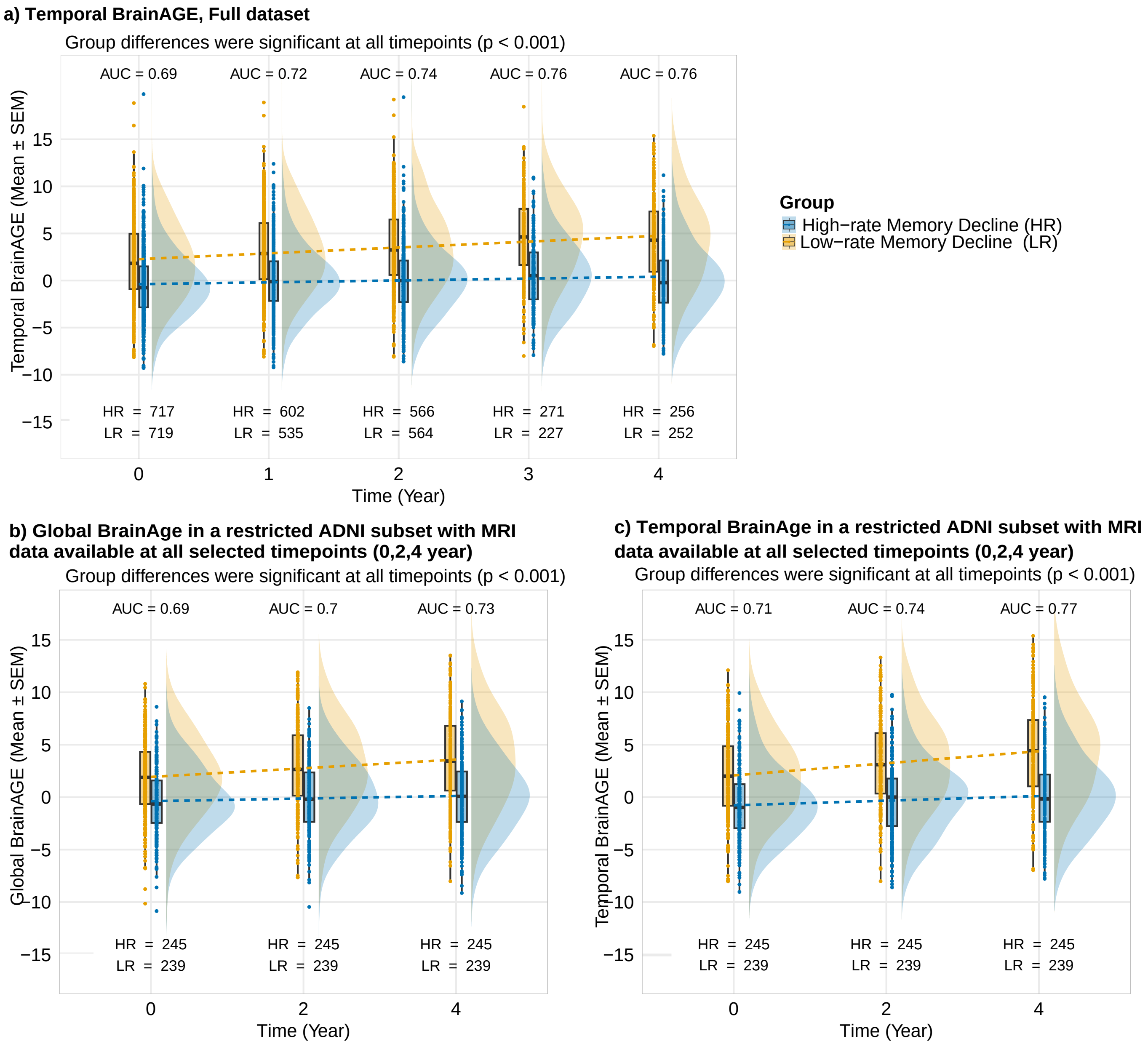


Figure S7: Longitudinal BrainAGE patterns in relation to memory decline in ADNI. (a) Temporal BrainAGE in participants with at least two MRI examinations (baseline and at least one follow-up within four years). (b, c) Global and Temporal BrainAGE, respectively, in a restricted subset with MRI data available at all selected time points. Dashed lines indicate group-level trends estimated using linear regression fitted separately for each group. Sample sizes and univariate AUC values are shown at each time point. These plots show higher BrainAGE values and steeper longitudinal increases in individuals with faster memory decline, with similar patterns observed in the restricted longitudinal subset.

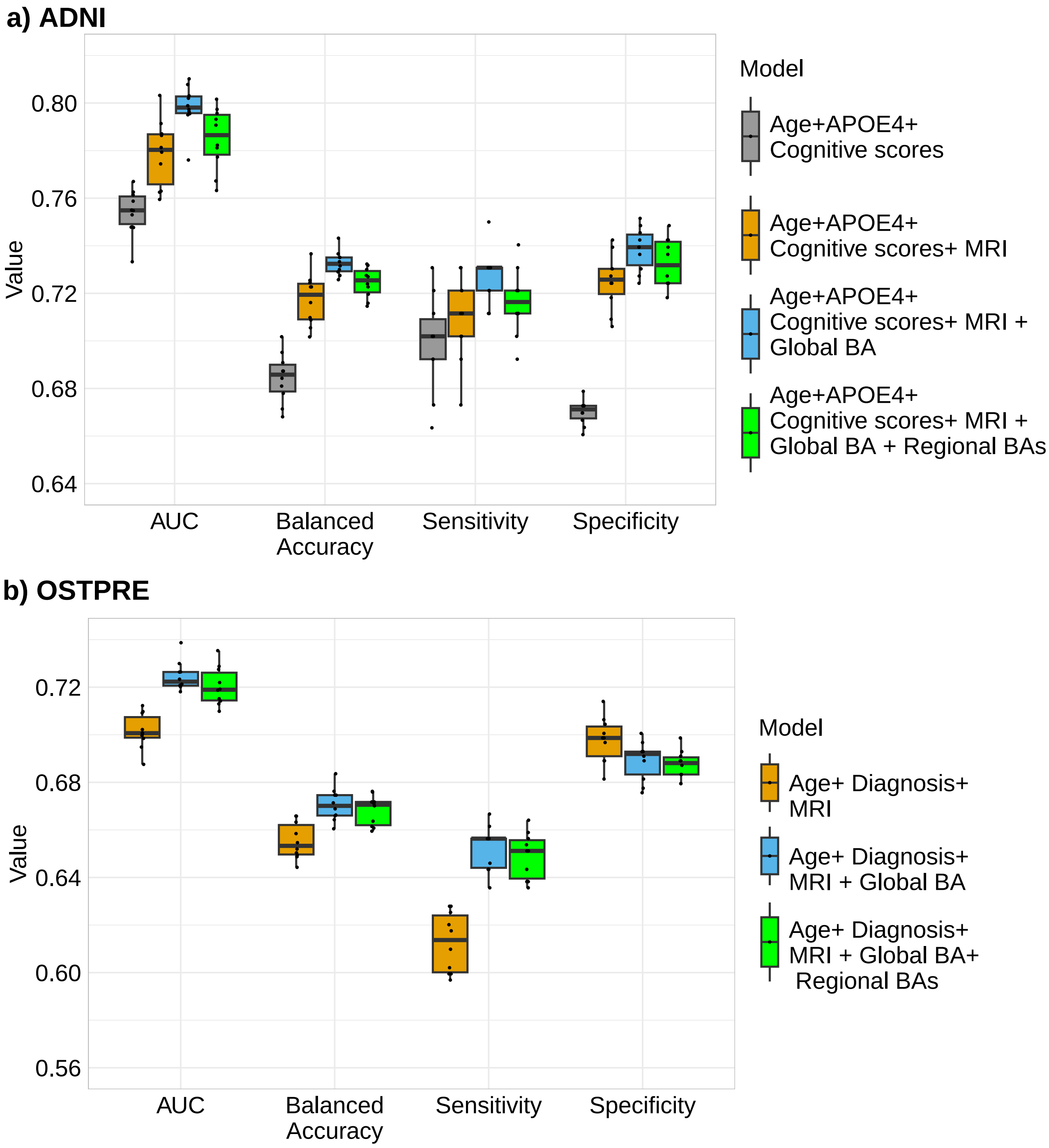


Figure S8: Distribution of predictive performance metrics across 10 repeated nested cross-validation runs for progression to MCI or dementia prediction models in the ADNI and OSTPRE cohorts. Boxplots summarize the distribution of the area under the receiver operating characteristic curve (AUC), balanced accuracy, sensitivity, and specificity for models with increasing information content. In ADNI, models included age, APOE4 status, baseline composite cognitive scores, the MRI-derived score, and BrainAGE measures. In OSTPRE, models included age, baseline cognitive status, the MRI-derived score, and BrainAGE measures, as APOE4 status and harmonized cognitive composite scores were unavailable. Inclusion of BrainAGE measures resulted in modest but consistent improvements in predictive performance across both cohorts.
